# Multi-ancestry Genome-Wide Association Study of Early Childhood Caries

**DOI:** 10.1101/2024.03.12.24303742

**Authors:** P Shrestha, M Graff, Y Gu, Y Wang, CL Avery, J Ginnis, MA Simancas-Pallares, AG Ferreira Zandoná, HS Ahn, KN Nguyen, DY Lin, JS Preisser, GD Slade, ML Marazita, KE North, K Divaris

## Abstract

Early childhood caries (ECC) is the most common non-communicable childhood disease. It is an important health problem with known environmental and social/behavioral influences that lacks evidence for specific associated genetic risk loci. To address this knowledge gap, we conducted a genome-wide association study of ECC in a multi-ancestry population of U.S. preschool-age children (n=6,103) participating in a community-based epidemiologic study of early childhood oral health. Calibrated examiners used ICDAS criteria to measure ECC with the primary trait using the dmfs index with decay classified as macroscopic enamel loss (ICDAS ≥3). We estimated heritability, concordance rates, and conducted genome-wide association analyses to estimate overall genetic effects; the effects stratified by sex, household water fluoride, and dietary sugar; and leveraged the combined gene/gene-environment effects using the 2-degree-of-freedom (2df) joint test. The common genetic variants explained 24% of the phenotypic variance (heritability) of the primary ECC trait and the concordance rate was higher with a higher degree of relatedness. We identified 21 novel non-overlapping genome-wide significant loci for ECC. Two loci, namely *RP11-856F16*.*2* (rs74606067) and *SLC41A3* (rs71327750) showed evidence of association with dental caries in external cohorts, namely the GLIDE consortium adult cohort (n=∼487,000) and the GLIDE pediatric cohort (n=19,000), respectively. The gene-based tests identified *TAAR6* as a genome-wide significant gene. Implicated genes have relevant biological functions including roles in tooth development and taste. These novel associations expand the genomics knowledge base for this common childhood disease and underscore the importance of accounting for sex and pertinent environmental exposures in genetic investigations of oral health.

## Introduction

Early childhood caries (ECC) is the most common non-communicable disease of childhood with a reported global prevalence of 46% (Kazeminia et al. 2020). It is an early-onset form of dental disease defined by the presence of one or more primary tooth surfaces with caries experience in a child under the age of six. Efforts to better understand, treat, and prevent this persistent disease, must include disentangling its social/behavioral and biological determinants and represent populations that experience high burdens of disease but may be underrepresented in research.

Dental caries is now understood as a complex dysbiotic disease resulting from the interplay between environmental and genetic etiologic factors (Divaris 2016). About a dozen genome-wide association studies (GWAS) of dental caries in children and adults have been reported; however, only two studies have interrogated the early-onset, severe form of disease that is captured in ECC (Borgio et al. 2021; Orlova et al. 2022). To date, 7 loci for caries in children (including those over the age of 6) have been reported, yet studies have been limited by small sample sizes, a focus on European populations (∼90%), heterogeneous phenotypic characterization, and wide age intervals.

As with most complex diseases, genetic effects on ECC may differ according to environmental exposures. Non-genetic factors that play an important role in the etiology of dental caries include sugar consumption, fluoride exposure, oral hygiene, the oral microbiome, and biological sex, among others. Indeed, genetic studies have demonstrated that accounting for environmental heterogeneity can aid the detection of genetic associations that may be under the radar in main-effects analyses alone (Aschard et al. 2010). Yet, there is a paucity of genetic studies on ECC, both overall and with consideration of environmental heterogeneity. To address this knowledge gap and add to the evidence base of genetic determinants of ECC, we carried out a GWAS leveraging potential gene-environment (GxE) interactions to identify genetic risk loci associated with ECC in a multi-ancestry population of preschool-age children.

## Methods

### Study population

The analytical sample comprised a multi-ethnic cohort of 6,103 preschool-age children participating in the ZOE 2.0 study in North Carolina, United States (Appendix Fig. 1) (Divaris et al. 2020). Approximately 48% of participants were non-Hispanic African Americans, 20% Hispanic Americans, 18% non-Hispanic Whites, among others, and 50% were females (Appendix Table 1). More information on study methodology can be found in the supplemental material (Appendix).

### Phenotypes

The primary quantitative phenotype (“cavitated decay” or d_3-6_mfs) was defined as the number of caries-affected tooth surfaces [with caries lesions considered at the International Caries Detection and Assessment System (ICDAS)≥3 threshold] (Pitts and Ekstrand 2013), missing or filled due to dental caries, i.e. the decayed-missing-filled surfaces (dmfs) index. Three secondary ECC traits considered were: a quantitative ECC dmfs index (“clinical decay” or d_1-6_mfs) that included early-stage caries lesions (i.e., both cavitated and non-cavitated lesions, ICDAS≥1) (Ginnis et al. 2019); and two binary ECC case status traits (i.e., dmfs>0) corresponding to the two quantitative traits defined above (Appendix Table 2).

### Genotyping and imputation

Saliva samples were collected using the DNA Genotek Oragene DNA-575 kit (DNA Genotek, Ottawa, Ontario, Canada). High-density genotyping of purified DNA was performed at the Center for Inherited Disorders Research (CIDR), at Johns Hopkins University, using the Infinium™ Global Diversity Array-8 v1.0 (Illumina, San Diego, CA, USA). Imputation was carried out at CIDR for 6,103 unique, genotyped study participants using the Trans-Omics for Precision Medicine (TOPMed) imputation server.

### Heritability estimates

Heritable variance (h^2^) of ECC attributable to all GWAS SNPs was estimated among 5,580 unrelated participants using Genome-wide Complex Trait Analysis (GCTA) using genotyped and high-quality imputed SNPs (R^2^>0.7); excluding SNPs with MAF<5%; and adjusting for age, sex, eight ancestry principal components and self-reported race/ethnicity (Yang et al. 2013).

### Concordance estimates

We estimated the concordance of ECC among 682 pairs of related individuals using Cohen’s *kappa* for the two categorical case statuses and intraclass correlation coefficients (ICC) for the two quantitative traits.

### Statistical analyses

The modelling considerations for phenotypes and accounting for complex study design is discussed in the supplemental material (Appendix notes). We used three approaches for the genome-wide association (GWA) testing (Fig. 1). Approach 1 (main discovery) was a GWAS in the entire study sample. We used linear and logistic mixed models, respectively for quantitative and binary traits assuming an additive genetic model. We adjusted each model for age, sex, race/ethnicity, first 8 ancestry principal components, sugary snacks/beverages, and fluoride content of household water (i.e., fixed effects) (Appendix Table 3). In the second approach (SNP_joint_), we investigated single variant associations with ECC testing the hypothesis that a variant has a main and/or interaction effect on ECC using a joint 2-degree-of-freedom (2df) test (Aschard et al. 2010). This approach leveraged potential gene-environment interaction effects in the development of ECC, accounting for the heterogeneity of genetic effects across different strata of interest, i.e. (a) sex, (b) daily between-meal consumption frequency of high (2 or more) versus low (0-1) sugary snacks and beverages, and (c) exposure to optimal (≥0.6ppm) versus sub-optimal level (<0.6ppm) of domestic water source fluoride. The third approach entailed stratified analyses, wherein the sample was split in two, for each of the three environmental exposures. For loci demonstrating genome-wide significant evidence of association in any of the previous analyses, we calculated the p-value for difference (P-difference) between the stratum-specific beta-coefficients of lead SNPs to screen for evidence of GxE interaction effects.

**Figure 1.**
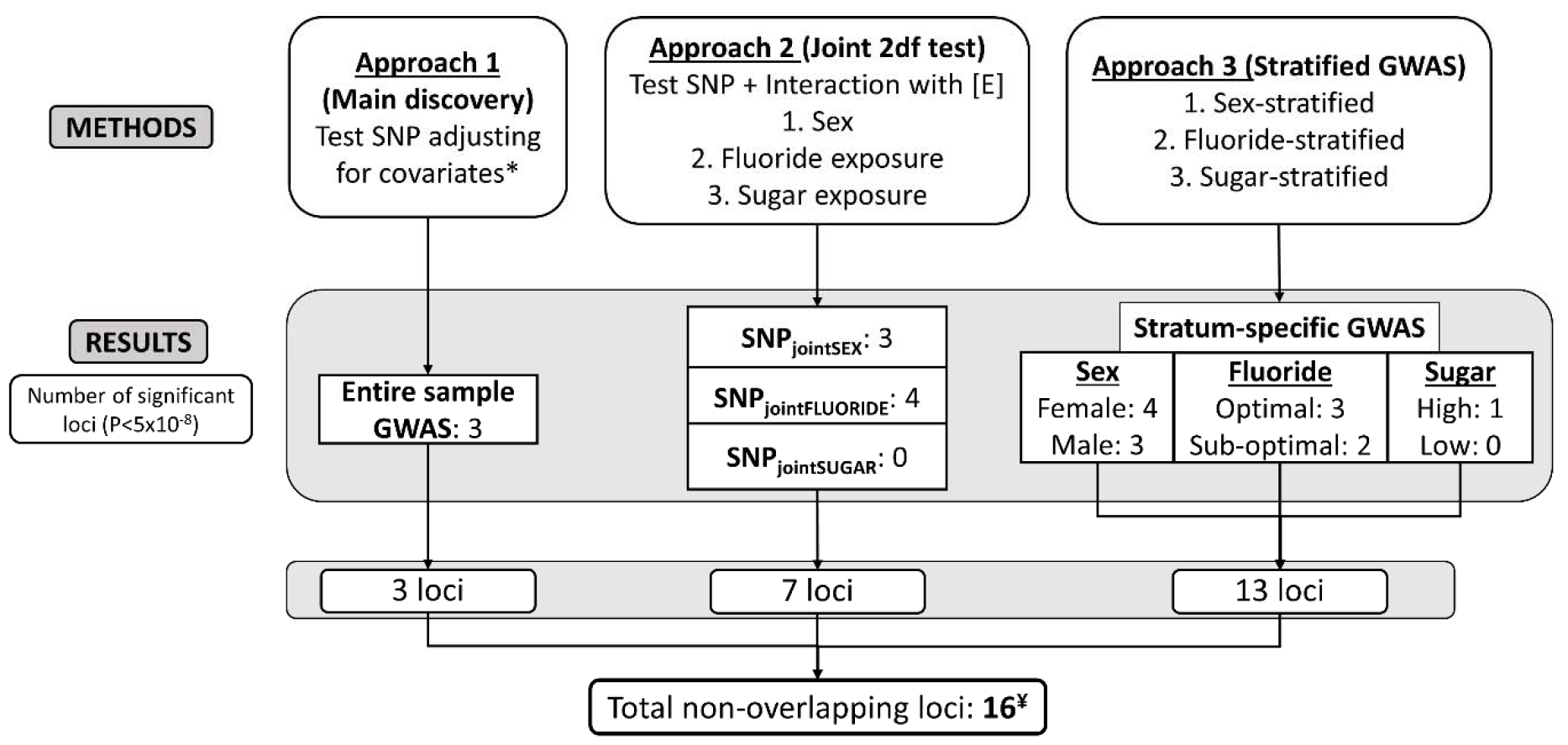
Summary of study design and results. In approach 1, we conducted a genome-wide association study (GWAS) in the entire sample. In Approach 2, we conducted a joint 2-degree-of-freedom test to test the main and interaction effects jointly accounting for interactions with 3 environmental exposures, i.e., the sex, fluoride exposure (fluoride content of household water), and sugar exposure (sugary snacks and beverages in-between meals). In Approach 3, we conducted stratified GWAS for the 3 dichotomous environmental exposures. ^*^Covariates in all association tests included age, sex, race/ethnicity, first 8 ancestry principal components, fluoride exposure, and sugar exposure. ^**¥**^Further, 5 loci were independently genome-wide significant for the secondary ECC traits (Appendix table 7). Abbreviations: SNP: Single Nucleotide Polymorphism, 2df: 2-degree-of-freedom, [E]: Environmental factor, GWAS: Genome-wide association study, P: p-value.

The R package EasyStrata was used to perform quality control (QC), generate Manhattan and quantile-quantile (Q-Q) plots, and conduct 1df, 2df, and P-difference tests (Winkler et al. 2015). We used SAIGE for all genetic association analyses, and accounted for relatedness using a genetic relationship matrix (Zhou et al. 2018). To identify genome-wide significant signals, we excluded variants with MAF<1%, R^2^<0.3 and excluded imputed SNPs with small effective sample sizes (effN<20 for combined and effN<40 for stratified analyses) resulting in the test of ∼14 million autosomal SNPs (Appendix Table 4). A multiple testing-corrected statistical significance criterion of P<5x10^−8^ was used for all analyses (Uffelmann et al. 2021). We reported genome-wide significant loci only if the lead SNP had effN≥100 to reduce likelihood of reporting spurious associations. The environmental exposure groups are described in the Appendix and the stratified baseline characteristics presented in Appendix Table 1.

### Generalization

We examined the summary estimates of the genome-wide statistically significant SNPs in our study for directional consistency and nominal statistical significance (P<0.05) in two genome-wide meta-analysis of dental caries conducted among children (Haworth et al. 2018) and adults (Shungin et al. 2019). We considered the variants fulfilling both these criteria as generalized.

### Functional annotation

We used FUMA GWAS (Functional Mapping and Annotation of Genome-Wide Association Studies) to facilitate functional annotation of single variant testing results (Watanabe et al. 2017). Additionally, we used FATHMM-XF, HaploReg, GTeX, GeneCards, and GWAS-catalog for further biological annotation.

### Gene-based test, gene-set analysis, and pathway enrichment test

MAGMA v1.6 was used to perform gene-centric analyses within the SNP2GENE process of FUMA. Furthermore, genes prioritized from SNP2GENE were tested in the GENE2FUNC process using hypergeometric tests to evaluate pathway enrichment in pre-defined gene sets from MsigDB, WikiPathways, and GWAS catalog.

## Results

### Heritability and trait concordance (Table 1)

We estimated that a quarter of variance in quantitative cavitated decay trait was explained by common GWAS SNPs (i.e., dmfs index based on the ICDAS≥3 criterion, h^2^=0.24, SE=0.07, P=9.8x10^−5^). A genome-wide interaction term for optimal fluoride exposure was statistically significant (P=2.9x10^−3^) and resulted in the variance explained increasing to 28% (h^2^=0.28, SE=0.07, P=3.7x10^−2^), a 17% relative increase. A weaker genome-wide interaction was found for sugary snacks consumption frequency (P=7.1x10^−2^). Additionally, we found higher concordance amongst participants with a higher level of relatedness; for example, concordance for the cavitated decay trait was 0.64 (95% CI=0.42-0.79) for monozygotic twins, 0.44 (95% CI=0.34-0.53) for first degree relatives and 0.13 (95% CI=0.03-0.23) for 2^nd^ and 3^rd^ degree relatives.

**Table 1:** SNP-based heritability estimates with and without the inclusion of Gene x Environment interaction terms with sugar exposure and fluoride level in household water among unrelated individuals. Concordance (*kappa* or ICC and corresponding 95% confidence intervals) of quantitative and binary ECC traits among pairs of related individuals at different levels of relatedness (assessed using the kinship coefficient).

|  | Quantitative (number of affected tooth surfaces) |  | Binary case status (any versus no affected tooth surfaces) |  |
| --- | --- | --- | --- | --- |
| Traits | Cavitated decay (d<sub>3-6</sub>-mfs)* | Clinical decay (d<sub>1-6</sub>mfs)** | Cavitated decay status | Clinical decay status |
| **Heritability estimates** |  |  |  |  |
| h<sup>2</sup> (SE) | 0.24 (0.07) | 0.25 (0.07) | 0.24 (0.06) | 0.16 (0.07) |
| p | 9.8x10<sup>-5</sup> | 5.8x10<sup>-5</sup> | 1.1x10<sup>-4</sup> | 5.6x10<sup>-3</sup> |
| +GxE (sugar) (SE) | 0.26 (0.07) | 0.24 (0.06) | 0.24 (0.07) | 0.29 (0.08) |
| p (G) | 6.9x10<sup>-5</sup> | 8.9x10<sup>-4</sup> | 5.1x10<sup>-4</sup> | 3.0 x10<sup>-1</sup> |
| p (GxE) | 7.1x10<sup>-2</sup> | 5.0x10<sup>-1</sup> | 4.0x10<sup>-1</sup> | 1.0x10<sup>-2</sup> |
| +GxE (fluoride) (SE) | 0.28 (0.07) | 0.24 (0.07) | 0.24 (0.07) | 0.16 (0.07) |
| p (G) | 3.7x10<sup>-2</sup> | 6.7x10<sup>-4</sup> | 1.3x10<sup>-2</sup> | 9.3x10<sup>-3</sup> |
| p (GxE) | 2.9x10<sup>-3</sup> | 4.1x10<sup>-1</sup> | 3.4x10<sup>-1</sup> | 2.5x10<sup>-1</sup> |
| **Concordance estimates** |  |  |  |  |
| Monozygotic twins (41 pairs) | 0.64 [0.42, 0.79] | 0.64 [0.42, 0.79] | 0.79 [0.39, 1.00] | 0.30 [0.00, 0.59] |
| 1st degree relatives (259 pairs) | 0.44 [0.34, 0.53] | 0.52 [0.43, 0.60] | 0.22 [0.03, 0.41] | 0.25 [0.13, 0.36] |
| 2nd & 3rd degree relatives (382 pairs) | 0.13 [0.03, 0.23] | 0.16 [0.06, 0.26] | 0.02 [-0.09, 0.13] | 0.20 [0.10, 0.30] |

□ all estimates are adjusted for age, sex, race/ethnicity and first 8 ancestry principal components;
\*analysis used age, sex, and race/ethnicity-model adjusted Pearson residuals; \*\*natural log-transformed count. Abbreviations: ECC: Early childhood caries; h<sup>2</sup>: heritability estimate; SE: Standard error; p: p-value; GxE: GxE interaction term; p(G): p-value for the variance of main genetic effect; p(GxE): p-value for variance of gene-environment interaction effect

**Table 2:**
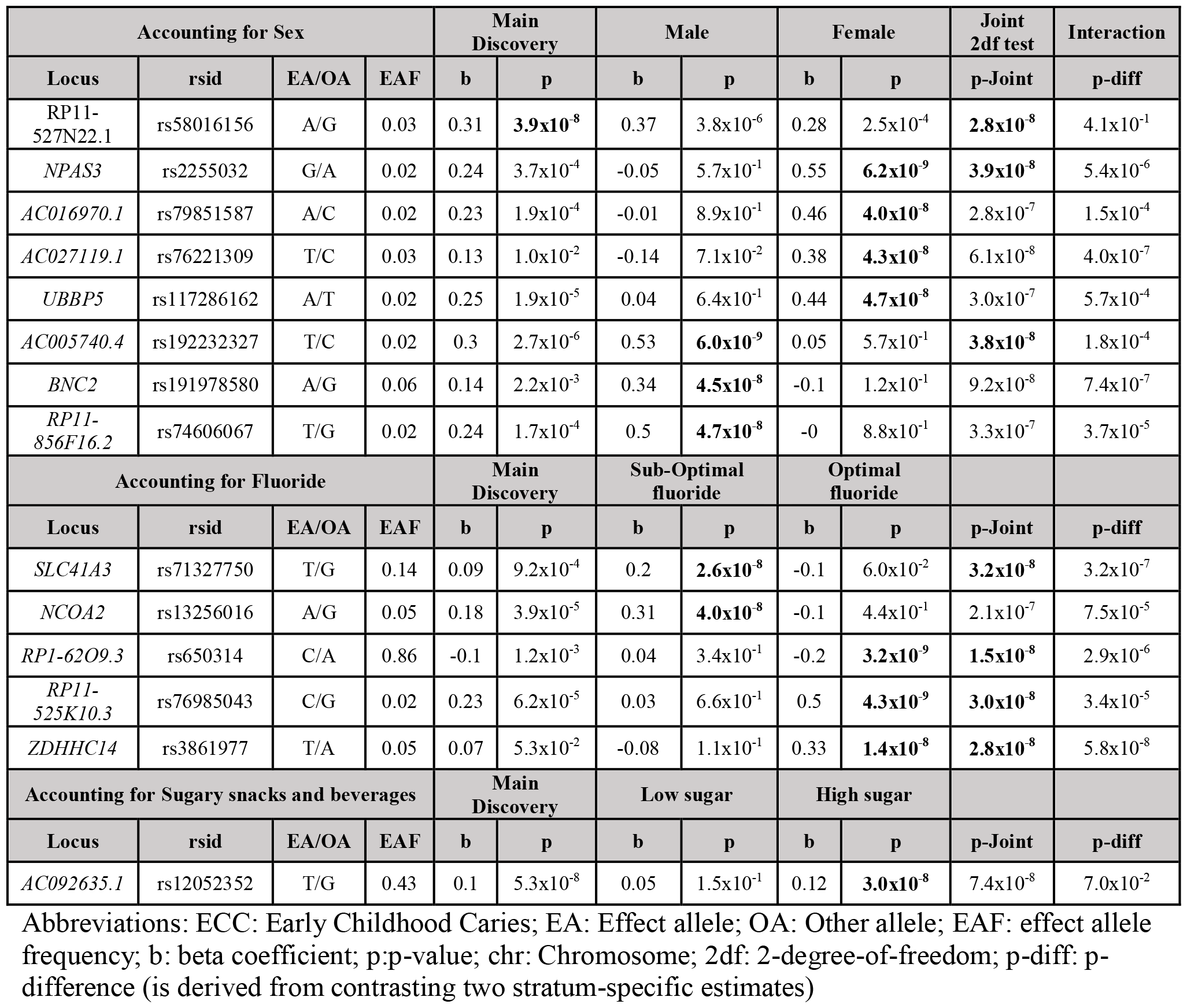
Summary of association results for the 14 loci that met genome-wide significance criteria (P<5x10^−8^) in analyses that accounted for heterogeneity by sex, fluoride, and sugar exposure, using 2 degree-of-freedom tests and stratified analyses.

| Accounting for Sex |  |  |  | Main Discovery |  | Male |  | Female |  | Joint 2df test | Interaction |
| --- | --- | --- | --- | --- | --- | --- | --- | --- | --- | --- | --- |
| Locus | rsid | EA/OA | EAF | b | p | b | p | b | p | p-Joint | p-diff |
| RP11-527N22.1 | rs58016156 | A/G | 0.03 | 0.31 | **$3.9 \times 10^{-8}$** | 0.37 | $3.8 \times 10^{-6}$ | 0.28 | $2.5 \times 10^{-4}$ | **$2.8 \times 10^{-8}$** | $4.1 \times 10^{-1}$ |
| NPAS3 | rs2255032 | G/A | 0.02 | 0.24 | $3.7 \times 10^{-4}$ | -0.05 | $5.7 \times 10^{-1}$ | 0.55 | **$6.2 \times 10^{-9}$** | **$3.9 \times 10^{-8}$** | $5.4 \times 10^{-6}$ |
| AC016970.1 | rs79851587 | A/C | 0.02 | 0.23 | $1.9 \times 10^{-4}$ | -0.01 | $8.9 \times 10^{-1}$ | 0.46 | **$4.0 \times 10^{-8}$** | $2.8 \times 10^{-7}$ | $1.5 \times 10^{-4}$ |
| AC027119.1 | rs76221309 | T/C | 0.03 | 0.13 | $1.0 \times 10^{-2}$ | -0.14 | $7.1 \times 10^{-2}$ | 0.38 | **$4.3 \times 10^{-8}$** | $6.1 \times 10^{-8}$ | $4.0 \times 10^{-7}$ |
| UBBP5 | rs117286162 | A/T | 0.02 | 0.25 | $1.9 \times 10^{-5}$ | 0.04 | $6.4 \times 10^{-1}$ | 0.44 | **$4.7 \times 10^{-8}$** | $3.0 \times 10^{-7}$ | $5.7 \times 10^{-4}$ |
| AC005740.4 | rs192232327 | T/C | 0.02 | 0.3 | $2.7 \times 10^{-6}$ | 0.53 | **$6.0 \times 10^{-9}$** | 0.05 | $5.7 \times 10^{-1}$ | **$3.8 \times 10^{-8}$** | $1.8 \times 10^{-4}$ |
| BNC2 | rs191978580 | A/G | 0.06 | 0.14 | $2.2 \times 10^{-3}$ | 0.34 | **$4.5 \times 10^{-8}$** | -0.1 | $1.2 \times 10^{-1}$ | $9.2 \times 10^{-8}$ | $7.4 \times 10^{-7}$ |
| RP11-856F16.2 | rs74606067 | T/G | 0.02 | 0.24 | $1.7 \times 10^{-4}$ | 0.5 | **$4.7 \times 10^{-8}$** | -0 | $8.8 \times 10^{-1}$ | $3.3 \times 10^{-7}$ | $3.7 \times 10^{-5}$ |
| Accounting for Fluoride |  |  |  | Main Discovery |  | Sub-Optimal fluoride |  | Optimal fluoride |  |  |  |
| Locus | rsid | EA/OA | EAF | b | p | b | p | b | p | p-Joint | p-diff |
| SLC41A3 | rs71327750 | T/G | 0.14 | 0.09 | $9.2 \times 10^{-4}$ | 0.2 | **$2.6 \times 10^{-8}$** | -0.1 | $6.0 \times 10^{-2}$ | **$3.2 \times 10^{-8}$** | $3.2 \times 10^{-7}$ |
| NCOA2 | rs13256016 | A/G | 0.05 | 0.18 | $3.9 \times 10^{-5}$ | 0.31 | **$4.0 \times 10^{-8}$** | -0.1 | $4.4 \times 10^{-1}$ | $2.1 \times 10^{-7}$ | $7.5 \times 10^{-5}$ |
| RP1-62O9.3 | rs650314 | C/A | 0.86 | -0.1 | $1.2 \times 10^{-3}$ | 0.04 | $3.4 \times 10^{-1}$ | -0.2 | **$3.2 \times 10^{-9}$** | **$1.5 \times 10^{-8}$** | $2.9 \times 10^{-6}$ |
| RP11-525K10.3 | rs76985043 | C/G | 0.02 | 0.23 | $6.2 \times 10^{-5}$ | 0.03 | $6.6 \times 10^{-1}$ | 0.5 | **$4.3 \times 10^{-9}$** | **$3.0 \times 10^{-8}$** | $3.4 \times 10^{-5}$ |
| ZDHHC14 | rs3861977 | T/A | 0.05 | 0.07 | $5.3 \times 10^{-2}$ | -0.08 | $1.1 \times 10^{-1}$ | 0.33 | **$1.4 \times 10^{-8}$** | **$2.8 \times 10^{-8}$** | $5.8 \times 10^{-8}$ |
| Accounting for Sugary snacks and beverages |  |  |  | Main Discovery |  | Low sugar |  | High sugar |  |  |  |
| Locus | rsid | EA/OA | EAF | b | p | b | p | b | p | p-Joint | p-diff |
| AC092635.1 | rs12052352 | T/G | 0.43 | 0.1 | $5.3 \times 10^{-8}$ | 0.05 | $1.5 \times 10^{-1}$ | 0.12 | **$3.0 \times 10^{-8}$** | $7.4 \times 10^{-8}$ | $7.0 \times 10^{-2}$ |

Abbreviations: ECC: Early Childhood Caries; EA: Effect allele; OA: Other allele; EAF: effect allele frequency; b: beta coefficient; p:p-value; chr: Chromosome; 2df: 2-degree-of-freedom; p-diff: p-difference (is derived from contrasting two stratum-specific estimates)

### Genome-wide association analysis

Population stratification was well-controlled as evidenced by genomic inflation factors (*lambdas* ranged between 0.98-1.02) (Appendix Table 5) and quantile-quantile (QQ) plots (Appendix Fig. 2). We identified 16 genome-wide significant loci for the primary quantitative ECC trait. We identified 3 loci in the main discovery GWAS (Fig. 2); 6 additional ones in the joint main effect/GxE approach, i.e., SNP_joint_ (2 for SNP_jointSEX_, 4 for SNP_jointFLUORIDE_); 5 additional ones in the sex-stratified, 1 in the fluoride-stratified and 1 in the sugar-stratified analysis (Fig. 1). We summarize the findings from approaches 2 and 3 in Table 2 (details in Appendix Tables 6-7; Manhattan plots in Appendix Fig. 3 and 4).

**Figure 2.**
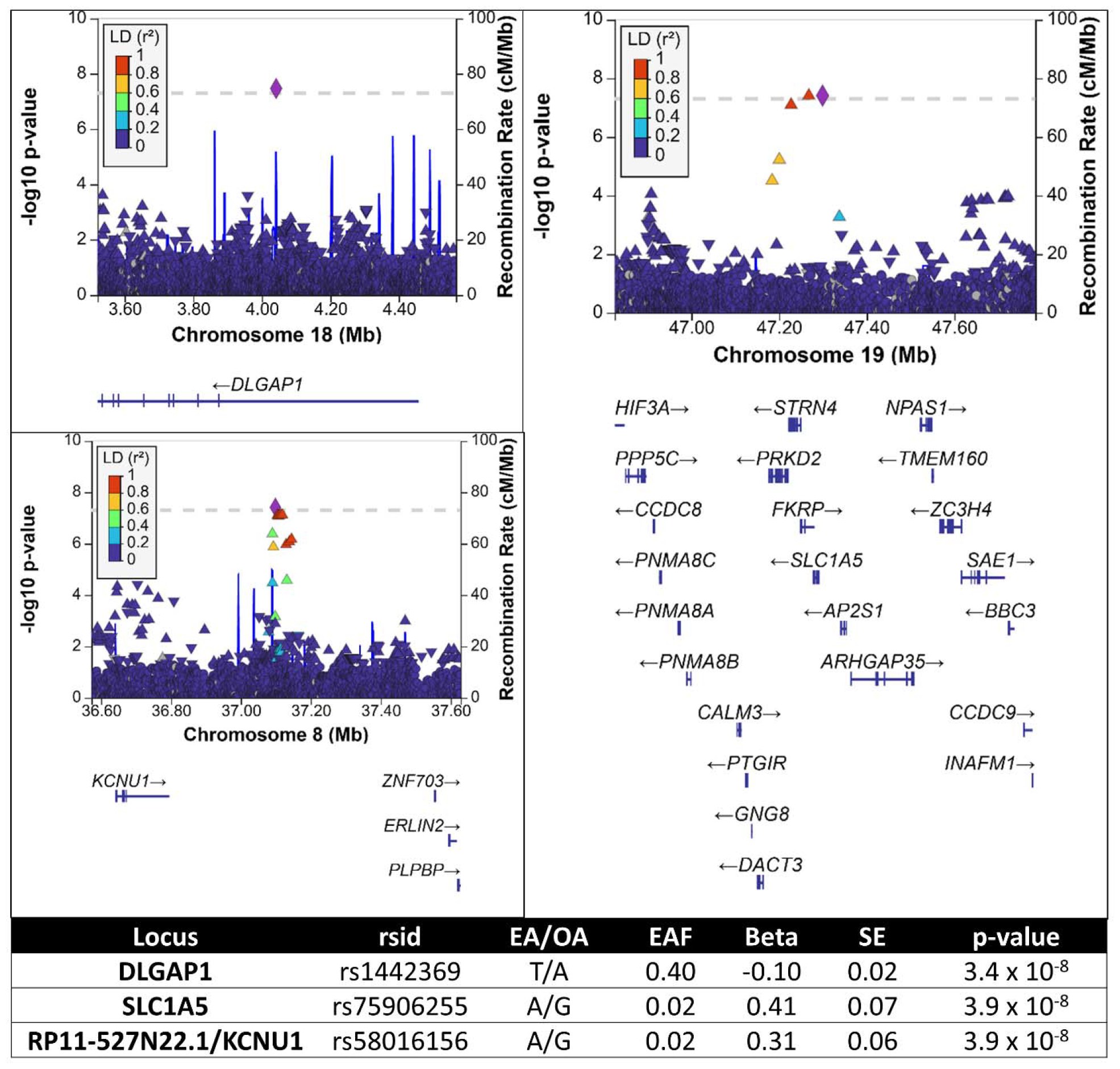
Regional association plots and summary of association results of the three genetic risk loci for ECC from the main discovery analysis (Approach 1) in a multi-ancestry population of preschool-age children. (a) DLGAP1 (b) SLC1A5 (c) KCNU1. Vertical axes illustrate association p-values on the –log10 scale, and horizontal axes represent chromosome positions. Purple diamonds denote the SNP with the strongest association signal (lead SNP) in the locus. Other SNPs in locus are colored based on their LD (all populations, 1000G data) with the lead SNP. **Abbreviations:** ECC: Early Childhood Caries; EA: Effect allele; OA: Other allele; EAF: effect allele frequency; n: sample size; EffN: Effective sample size; b: beta coefficient; SE: standard error; chr: Chromosome; pos: Position; p: p-value

**Figure 3.**
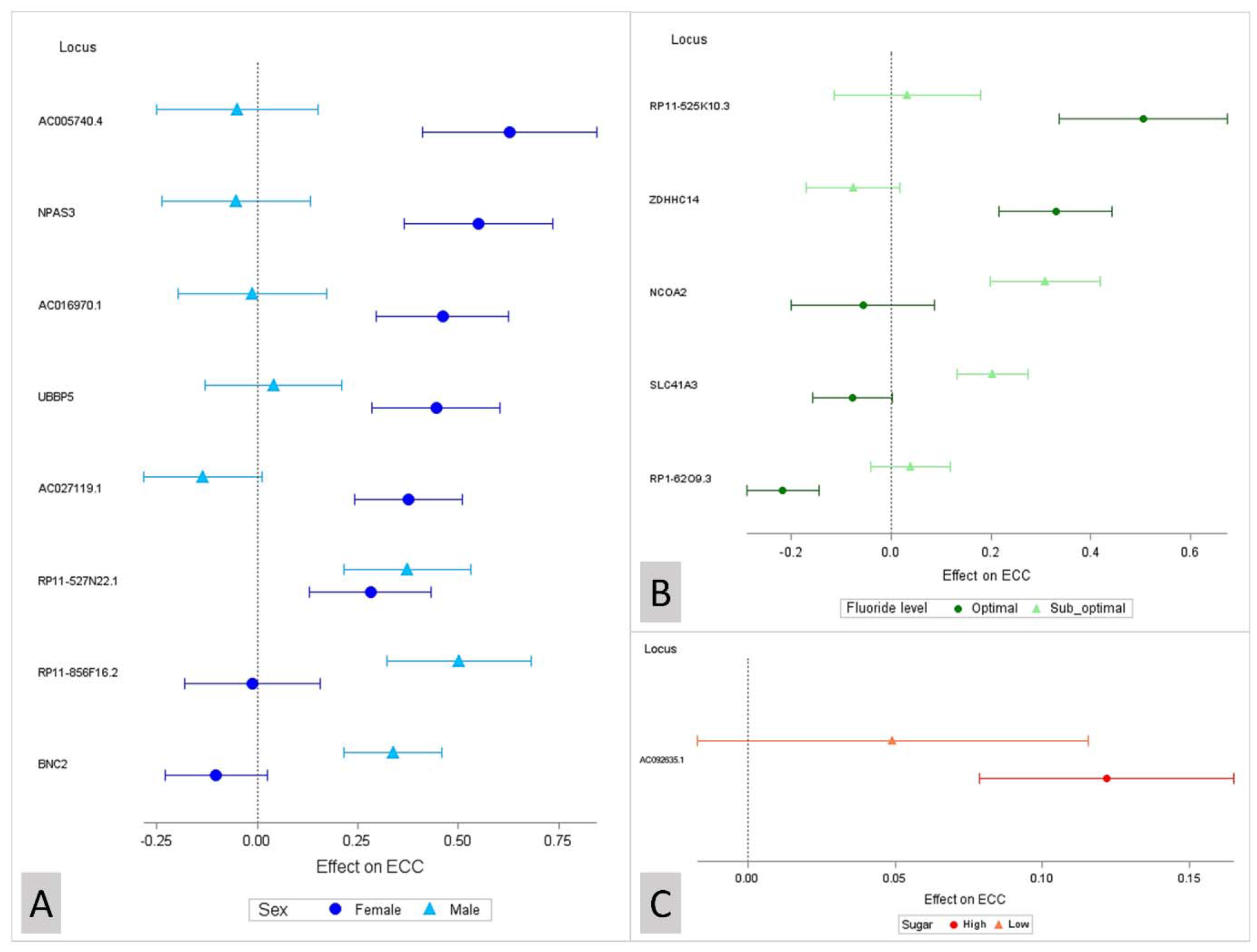
Forest plots demonstrating heterogeneity of genetic effect due to (A) Sex, (B) Fluoride exposure, and (C) Sugary snacks/beverage consumption among top loci identified in the three stratified GWASs (Approach 3). The triangles and circles represent the effect estimate of the loci for ECC and the error bars represent the 95% confidence interval. The loci are labelled as the nearest gene and ordered by the greater magnitude of association among females, optimal-fluoride level stratum, and the high-sugar stratum, respectively

### Discovery GWAS

In the main discovery analysis (Appendix Table 8) we identified 3 genome-wide significant loci on chromosomes 8 (lead SNP: rs58016156), 18 (rs1442369), and 19 (rs75906255). Rs1442369 (effect allele frequency (EAF) [T]: 0.40, P=3.4x10^−8^, beta=-0.10), is a variant intronic to *DLGAP1*. Rs75906255 (EAF[A]: 0.02, P=3.9x10^−8^, beta=0.41) is adjacent (∼7Kb) to *SLC1A5* and in LD with potentially functional variants rs77394147 (R^2^=0.93, RegulomeDB score [RDB]: 2b) and rs76308698 (R^2^=0.80, CADD score [CADD]: 10.4). Rs58016156 (EAF[A]: 0.02, P=3.9x10^−8^, beta=0.31), is an intergenic variant adjacent to (∼89Kb) *RP11-527N22*.*1* and 305Kb upstream of *KCNU1* gene, which is part of the sweet taste signaling pathway.

### SNP_joint_ tests

Accounting for sex led to identification of two additional loci on chromosomes 14 (rs2255032, EAF[G]: 0.03; *NPAS3*) and 5 (rs192232327, EAF[T]: 0.03; *AC005740*.*4*) (Table 2). Both loci were also genome-wide significant in male and female-stratified analyses, respectively. Rs2255032 (intronic to *NPAS3)* has a CADD score of 11.7 and is in LD with a potentially functional variant rs74775070 (R^2^=0.67, CADD score: 17.4).

Accounting for optimal fluoride in household water led to the identification of 4 additional loci, all of which were also genome-wide significant in fluoride-stratified analyses: chromosomes 3 (rs71327750, EAF[T]: 0.14; *SLC41A3*), 17 (rs650314, EAF[T]: 0.86; *RP1-62O9*.*3*), 16 (rs76985043, EAF[C]: 0.02, *RP11-525K10*.*3*), and 6 (rs3861977, EAF[T]: 0.05; *ZDHHC14*). Rs71327750 is intronic to *SLC41A3* and in LD with multiple potential functional variants, i.e., rs1077620 (R^2^=0.74, RDB: 2b); rs6796610 (R^2^=0.65, RDB: 1f); rs13100420 (R^2^=0.64, CADD: 12.6); rs4314124 (R^2^=0.65, RDB: 1f); rs35839813 (R^2^=0.61, RDB: 2b). The less common variant, rs76985043 in *RP11-525K10*.*3* is also in LD with multiple potentially functional variants [rs77285614 (R^2^=0.81, CADD: 10.1); rs76805928 (R^2^=0.73, CADD: 12.0); and rs79906923 (R^2^=0.75, RDB: 2b].

Accounting for sugary snacks and beverages in-between meals did not lead to the identification of any additional loci. The strongest signal was produced by a chromosome 2 locus (rs12052352, EAF[T]: 0.43; *AC092635*.1) which emerged as genome-wide significant in sugar-stratified analyses.

### Stratified GWAS

The sex-stratified analysis revealed 5 additional genome-wide significant loci. Three emerged only among females, on chromosomes 3 [(rs79851587; *AC016970*.*1*) and (rs76221309; *AC027119*.*1*)] and 13 (rs117286162; *UBBP5*), and two emerged only among males, on chromosomes 9 (rs191978580; *BNC2*) and 11 (rs74606067; *RP11-856F16*.*2*). Fluoride-stratified analysis revealed one additional genome-wide significant locus on chromosome 8 that emerged in the sub-optimal fluoride stratum. The lead SNP is rs13256016 intronic to *NCOA2*, and in LD with potentially functional variants: rs13269274 (R^2^=0.72, CADD: 13.7) and rs11784848 (R^2^=0.71, RDB: 2b). Finally, the sugar-stratified GWAS identified a chromosome 2 genome-wide significant locus (*AC092635*.*1*; rs12052352, EAF[T]: 0.43) in the high-sugar stratum. Upon comparison of stratum-specific estimates, we found that most of identified signals remained significantly different after a Bonferroni correction (Fig. 3).

### Secondary ECC traits (Appendix Table 7)

We discovered 5 genome-wide significant loci for the three secondary ECC traits. In the female stratum-specific analysis for the sensitive dmfs trait (including early-stage lesions), we identified a genome-wide significant signal led by rs4899701 (EAF[T]: 0.31, P=3.6x10^−8^, beta=-0.17) located 10Kb downstream of *NRXN3* (Neurexin 3) locus. This gene has been associated with obesity, autism spectrum disorder, schizophrenia, and alcohol dependence (Heard-Costa et al. 2009; Tromp et al. 2021). For the same trait in the optimal fluoride stratum-specific analysis, rs12420136 (EAF[G]: 0.04, P=2.5x10^−8^, beta=0.39), an intronic variant in the locus *MACROD1* was genome-wide significant. A gene-sex interaction effect has been reported for the *MACROD1* gene for early-onset periodontitis (Freitag-Wolf et al. 2021). Furthermore, rs11231965, an intergenic variant near (∼147Kb) *CTD-2555I5*.*1* was genome-wide significant in the joint 2df test for the sugar-stratified analysis and in the high-sugar stratum-specific analysis (EAF[G]: 0.11, P=7.8x10^−9^, beta=-0.21, P-joint=3.0x10^−8^). For the binary cavitated decay status trait, we identified one genome-wide significant locus in the female stratum (*RP11-215I16*.*1;* rs200747282) and one in the male stratum (*RP11-933H2*.*4/NUDT16P1;* rs35487488).

### Cross-trait and cross-test relevance of identified loci

We inspected all 21 loci’s estimates of association across all analyses and traits interrogated in this study (Appendix Table 9). Seven loci were genome-wide significant in two different analyses; e.g., rs58016156 in the *RP11-527N22*.*1/KCNU1* locus in the main GWAS and the joint test for the sex-stratified analysis. Of note, *SLC1A5* and *RP11-527N22*.*1/KCNU1* were associated with ECC at a suggestive significance level (p<5x10^−6^) in 7 and 6 different analyses, respectively.

### Generalization in external cohorts of children and adults

Among the 16 genome-wide significant signals for the primary ECC trait, 2 met the criteria of directional consistency and nominal significance (p<0.05) in the external cohorts. Rs74606067 (*RP11-856F16*.*2*), generalized in the GLIDE-adults cohort (EAF[A]: 0.53, P=0.03, N=285,246 and EAF[T]: 0.06, P=0.01, N=285,248). Rs71327750 (*SLC41A3*), generalized in the GLIDE-children cohort examining caries in primary teeth (EAF[T]: 0.20, P=0.02, N=18,994) (Appendix Tables 10-12).

We evaluated the previously published risk loci (P<5x10^−8^) in our study’s results (Appendix Table 13). Rs1122171 (*C5orf66*), a variant associated with caries in adults (Shungin et al. 2019), was nominally significant and had a directionally consistent estimate of association in our results for the primary trait. We did not find any associations listed for the statistically significant variants from our study or their proxies (R^2^≥0.8) in the GWAS-Catalog.

### Functional annotation

We queried multiple annotation tools to determine the functional significance of the total 21 identified loci (Appendix Table 14). We considered and summarized all protein-coding genes within ∼250Kb of the independently significant variants (Research data). We identified several genes near genome-wide significant loci with potential roles in the development of dental tissues. For example, *SPRY4* near rs192232327, antagonizes fibroblast growth factor (Klein et al. 2006), which is important at different stages of tooth development (Thesleff 2006) and *PHOSPHO1* near rs650314 is involved in dental tissue mineralization (Pandya et al. 2017).

### Gene-based tests and gene-set analyses

We identified 1 genome-wide significant gene for the primary ECC trait, *TAAR6* (Appendix Table 15 and Appendix Fig. 5) and 11 genome-wide significant gene-sets for primary and secondary ECC traits (Appendix Table 16). These included “taste receptor activity”, a biologically pertinent gene set given the necessity of fermentable carbohydrates in the mechanistic pathway underlying dental caries development and the important role taste plays in dietary preferences.

### Pathway enrichment tests

Our analyses identified several enriched curated gene sets, including the positional gene set chr20q12 (Appendix Table 17); *PLCG1, ZHX3, LPIN3, EMILIN3*, and *CHD6* were the prioritized genes overlapping with this gene set. Imhof and colleagues have demonstrated an upregulation of *EMILIN-3* in dentin caries lesions (Imhof et al. 2020).

## Discussion

In this GWAS among a well-characterized, community-based, multi-ethnic cohort of preschool-age children we leveraged approaches accounting for gene-environment interactions and identified 21 novel risk loci for ECC. We demonstrate that a quarter of variance in this common-complex childhood disease can be explained by common genetic variation and that the joint consideration of established environmental factors like sugar consumption and fluoride exposure increases phenotypic variance explained. Two of the identified signals generalized in external, independent populations, and several genes harbored in these loci have plausible biological roles in the pathogenesis of ECC and are promising targets for future investigations.

The most prominent novel identified loci included *DLGAP1, SLC1A5*, and *KCNU1*. Rs1442369 and rs75906255, the lead variants in the *DLGAP1* and *SLC1A5* locus, respectively, showed relatively consistent evidence of association in all stratified analyses. The *KCNU1* locus (rs58016156) is related to the sweet taste signaling pathway (Safran et al. 2021). The 6 previously published GWASs of childhood caries and 2 pilot studies (Appendix Table 18) reported a total 7 genome-wide statistically significant SNPs. None of these generalized in our results. The sample sizes for these earlier studies were modest, with the exception of a meta-analysis (n=19,003) (Haworth et al. 2018). Furthermore, five of these studies used binary case statuses as analytical endpoints. Thus, the sample size of ∼6,000 and the detailed phenotypic characterization enabling inquiry of quantitative traits at different detection thresholds is a relative improvement. Moreover, the inclusion of traditionally underrepresented racial/ethnic backgrounds addresses some equity issue in oral and genetic research and has been shown to confer analytical advantages like increase in power, efficiency in replication, and refinement of genetic association signals across ancestral populations (Agler and Divaris 2020; Lin et al. 2021). Another strength of the study is the narrow age range (3-5 years), which reduces the likelihood of physiologic tooth exfoliation.

Among biological factors, potential differential effects of relevant genes between sexes may partly explain the difference in caries experience (Lukacs and Largaespada 2006) as has been reported for other traits (Randall et al. 2013). To our knowledge, only one study has presented evidence of gene-sex interactions in dental caries comparing sex-specific heritability estimates and between-sex genetic correlations (Shaffer et al. 2015). Our study, leveraging sex-heterogeneity via joint tests resulted in 2 additional independent signals. One of these loci harbors *SPRY4*, a gene important for dental development, antagonist of fibroblast growth factor (FGF) and the other receptor tyrosine kinase signaling, expressed in the mesenchyme of the tooth germs (Klein et al. 2006; Thesleff 2006). Interestingly, a suggestive genome-wide association of *SPRY4* with erosive tooth wear was recently reported (Alaraudanjoki et al. 2019).

Among the 4 signals significant in the joint tests for fluoride-stratified analysis, rs650314 is near (∼25Kb) *PHOSPHO1*. This gene activates pyrophosphate activity, is involved in bone mineralization and maturation, plays a role in mineralization of tooth enamel (McKee et al. 2013; Pandya et al. 2017), and is associated with childhood hypophosphatasia (Reibel et al. 2009).

*MTRR*, near the *LOC729506* locus was identified in the female-specific GWAS for the cavitated decay binary trait and has been associated with ECC and being underweight in a candidate gene study (Antunes et al. 2017). *BNC2* and *AMOTL1*, are loci identified in the male-specific GWAS and have been associated with orofacial clefting (Chernus et al. 2018; Strong et al. 2023).

In conclusion, we have identified 21 candidate risk loci associated with ECC using discovery and stratified analyses. Acknowledging the study’s limitations, significant loci harboring *SPRY4* and *PHOSPHO1*, with roles in tooth development, and *MTRR*, previously associated with caries, are plausible candidates for ECC. The finding of heterogeneity of allelic effect between sexes and across different levels of sugary snacks/beverages and fluoride level in water demonstrates a key role of gene-environment interactions in the biology of dental caries and bears importance to future frameworks for risk profiling, prevention and treatment tailoring, and the application in precision dentistry. Validation of these findings in future investigations, in different populations, utilizing similar analytical frameworks, and utilizing mechanistic studies is warranted to establish the identified loci as replicable and causal.

## Supporting information

Research Data Shrestha

## Data Availability

All data produced in the present study are available upon reasonable request to the authors

https://data.bris.ac.uk/data/dataset/2j2rqgzedxlq02oqbb4vmycnc2

https://data.bris.ac.uk/data/dataset/108de7fd6f8ff188ebb9ac07fe77bfb5

## Data Availability

Genotype and phenotype data for ZOE 2.0 are publicly available as part of dbGaP accession phs002232.v1.p1 “TOPDECC-Trans-omics for Precision Dentistry and Early Childhood Caries: Genome-Wide Genotyping (CIDR) and Microbiome in the ZOE 2.0 Study”. Genomic summary results from the GLIDE adults study are available via: https://data.bris.ac.uk/data/dataset/2j2rqgzedxlq02oqbb4vmycnc2 and GLIDE-children via: https://data.bris.ac.uk/data/dataset/108de7fd6f8ff188ebb9ac07fe77bfb5.

## Acknowledgements

This work was supported by research grants from the National Institutes of Health: National Institute for Dental and Craniofacial Research U01DE025046 (K.D., A.G.F.Z, D.Y.L, J.S.P, G.D.S., K.E.N) and National Human Genome Research Institute X01HG010871 (K.D.). The content is solely the responsibility of the authors and does not necessarily represent the official views of the funders.

The authors thank CIDR investigators and staff at Johns Hopkins University for carrying out genotyping and imputation for the project with support from a resource-allocation grant NIH/NIHGR X01-HG010871; Dr. Patricia V. Basta and her team at the UNC-Chapel Hill Biospecimen Processing facility for the accessioning, storage, and disbursement of the saliva and extracted nucleic acid samples in the ZOE studies; all study participants and their families for their contributions.

## Author Contributions

Shrestha P, contributed to data analyses and annotation, results interpretation, drafted and critically revised the paper. Graff M, contributed to data analyses and annotation, results interpretation, and critically revised the paper. Gu Y, contributed to data analyses and annotation, and critically revised the paper. Wang Y, contributed to data analyses and annotation, and critically revised the paper. Avery CL, contributed to results interpretation, and critically revised the paper. Ginnis J, contributed to data collection, and critically revised the paper. Simancas-Pallares MA, contributed to data collection, and critically revised the paper. Ferreira Zandoná AG, contributed to study design, and critically revised the paper. Ahn HS, contributed to data analyses and annotation, and critically revised the paper. Nguyen KN, contributed to data analyses and annotation, and critically revised the paper. Lin DY, contributed to results interpretation, and critically revised the paper. Preisser JS, contributed to study design, results interpretation, and critically revised the paper. Slade GD, contributed to study design, and critically revised the paper. Marazita ML, contributed to results interpretation, and critically revised the paper. North KE, contributed to conceptualization, study design, supervision, results interpretation, and critically revised the paper. Divaris K, contributed to conceptualization, study design, supervision, data collection, results interpretation, and critically revised the paper. K.D. and K.E.N. contributed equally. All authors gave their final approval and agree to be accountable for all aspects of the work.

