## Appendix for "Multi-ancestry Genome-Wide Association Study of Early Childhood Caries"

Poojan Shrestha, Mariaelisa Graff, Yu Gu, Yujie Wang, Christy L. Avery, Jeannie Ginnis, Miguel A. Simancas-Pallares, Andrea G. Ferreira Zandoná, Sean H. Ahn, Ken N. Nguyen, Danyu Y. Lin, John S. Preisser, Gary D. Slade, Mary L. Marazita, Kari E. North*, Kimon Divaris*

**APPENDIX NOTES**

**Study design and population**. ZOE 2.0 is a cross-sectional, community-based, multiethnic study conducted in North Carolina, United States. Between 2016 and 2019, 8,059 children ages 3-5 enrolled in public preschools (Head Start) were enrolled in this IRB-approved (UNC-Chapel Hill #14-1992) study. Written informed consent was obtained from parents or legal guardians of all participants. Details of the study design, methods, and characteristics of the cohort have been reported in Divaris et al. 2020 (Divaris et al. 2020). The flowchart depicting recruitment, enrollment, exclusions, and the genotyped population eligible for analysis (n=6,144) is presented in Appendix Fig. 1. Based on kinship coefficients, the genotyped population comprised 41 monozygotic (MZ) pairs, 259 full-sibling pairs and 382 pairs of second- and third-degree relatives (Appendix Fig. 6). One MZ twin in each pair was excluded resulting in a sample of 6,103. The sample included 50% females with a mean age of 53 months (Appendix Table 1). Among the 6,103 children, clinical data for ECC were missing for 28 participants (final analysis sample n=6,075).

**Population stratification.** Principal component analysis (PCA) was used to examine the sample’s population structure and to produce sample eigenvectors (EV) to adjust for population stratification in further analyses (Patterson et al. 2006). We also estimated continental-ancestry proportions with a model-based analysis using ADMIXTURE software (Alexander et al. 2009). A supervised ADMIXTURE analysis was performed on 5,580 unrelated study participants combined with 1000 Genomes reference samples under the assumption of three or four ancestral populations (African, American Admixed, European for k=3; and Asian for k=4).

Forty-eight percent of participants were non-Hispanic African Americans, followed by Hispanic Americans representing 20% and non-Hispanic Whites with approximately 18%. About 10% reported more than one race and 2% were American Indians or Alaskan Natives (Appendix Table 1). Appendix Fig. 7 presents the distribution of the first two ancestry principal components illustrating the considerable admixture of the study population. The continental-ancestry proportions (estimated using a four ancestral population assumption) further demonstrate the presence of admixture in our study population (Appendix Fig. 8).

**Phenotypes.** Clinical examinations were carried out by trained and calibrated examiner-recorder pairs using modified International Caries Detection and Assessment System (ICDAS) visual criteria to record tooth surface-level caries experience (Pitts and Ekstrand 2013). All examiners were calibrated for intra- and inter-examiner reliability with weighed *kappa* thresholds of 0.65 and 0.75, respectively (Ginnis et al. 2019). The primary caries experience trait was the number of tooth surfaces in an individual that are decayed (cavitated, ICDAS≥3), missing, or filled due to caries and is commonly referred to as the dmfs index (range: 0-88). We refer to this quantitative trait as “cavitated decay” or “d_3-6_mfs”).

Secondary analyses used a more inclusive classification of the decayed component of the dmfs index that enumerated early-stage (i.e., non-cavitated) caries lesions, i.e. ICDAS 1 and 2, referred to as “clinical decay” or “d_1-6_mfs”. The rationale was that in young children a considerable proportion of caries experience is represented by early stage lesions (Divaris et al. 2020). Additionally, we considered binary definitions of caries status (dmfs>0) for both cavitated decay and clinical decay. The rationale for this is that there is interest in analysis comparing caries-affected versus caries-free children (Appendix Table 2).

**Genotype data and Quality control.** Among participants with clinical data (n=6,404), saliva samples were collected using the DNA Genotek Oragene DNA-575 kit (DNA Genotek, Ottawa, Ontario, Canada) from 6,369 participants. DNA was extracted from the 6,262 samples meeting quality and quantity criteria for genotyping, and the quality was assessed using procedures detailed in the study’s genomics analysis protocol (Agler et al. 2019). The purified DNA material was carried forward to high-density genotyping at the Center for Inherited Disorders Research (CIDR), at Johns Hopkins University, using the Infinium™ Global Diversity Array-8 v1.0 (Illumina, San Diego, CA, USA). Rigorous quality assurance/quality control (QA/QC) procedures were performed at CIDR as previously described (Appendix Fig. 9 and Appendix Table 4) (Laurie et al. 2010). QC metrics used to filter SNPs included CIDR technical failures, missing call rates (≥2%), >1 discordant calls in 62 study duplicates, >1 Mendelian errors, Hardy-Weinberg equilibrium (p<8x10^-4^), sex difference in allele frequency (≥0.2) or heterozygosity (>0.3) for autosomes/XY, and positional duplicates. A total of 6,144 study samples were genotyped after 118 samples failed QC checks with the release of 1,821,357 SNPs.

**Imputation**. Imputation was carried out at CIDR for 6,103 unique, genotyped study participants using the Trans-Omics for Precision Medicine (TOPMed) imputation server. One of each of the 41 MZ twins were kept in the imputation process. The genotyped data were pre-phased using Eagle 2 (V2.4). Minimac4 software (v1.5.7) was used to impute unobserved genotypes to the TOPMed imputation reference panel (Fuchsberger et al. 2015; Das et al. 2016; Taliun et al. 2021). A total of 306,740,382 SNPs were imputed, with 143,332,340 (46.7%) non-monomorphic variants (Appendix Table 4). The quality of imputation was overall high, with the average R^2^ scores (measure of confidence in imputed dosages) for SNPs with minor allele frequency (MAF)>1% ranging between 0.973 to 0.988.

**Heritability estimates.** Heritable variance (h^2^) of ECC attributable to human genome was estimated among 5,580 unrelated participants using Genome-wide Complex Trait Analysis (GCTA) using genotyped and high-quality imputed SNPs (R^2^>0.7); excluding SNPs with MAF<5%; and adjusting for age, sex, eight ancestry principal components and self-reported race/ethnicity(Yang et al. 2013). GCTA is a two-step method; the first being estimation of a genetic relationship matrix (GRM) using all available SNPs for all individuals. In the second step, the GRM is carried forward to a restricted maximum likelihood analysis to estimate the proportion of variance explained by all SNPs.

**Concordance estimates.** We estimated the concordance of ECC traits among 682 pairs of related individuals using Cohen’s *kappa* for the two categorical case statuses and intraclass correlation coefficients (ICC) for the two quantitative traits.

**Statistical analyses.**

Modelling considerations for phenotypes**.** The distributions of both the cavitated decay (primary trait) and the clinical decay traits are right-skewed (Appendix Fig. 10). Furthermore, the distribution of the cavitated decay trait is zero-inflated. To examine model fits using these traits, we used linear regression models that included age (in months), sex, and race/ethnicity as covariates. We compared the diagnostic plots for the clinical decay trait and the log-transformed [log (trait+1)] clinical decay trait (Appendix Fig. 11 and 12). These diagnostic plots showed that the log transformation of the clinical decay trait presented a better model fit. Hence, we used the log-transformed form of the clinical decay trait for all genetic association analyses (Appendix Table 3).

To account for the zero inflation of the cavitated decay trait, we regressed this trait on age, sex, and race/ethnicity and produced Pearson residuals using a zero-inflated negative binomial (ZINB) model. These residuals were used as response variables in the genome-wide association analysis. For stratified genetic analyses, Pearson residuals were obtained for each stratum of the environmental exposure. We further compared the GWAS results produced using the Pearson residuals with the ones produced using the inverse normalized Pearson residuals. Genomic inflation factors (i.e., lambdas) and Q-Q plots were comparable for the discovery GWAS. However, when comparing sex-stratified GWA analyses using the two trait transformations, we noted considerable deflation in Q-Q plots and lambdas for the inverse normalized form. Given these comparisons and to avoid any loss of statistical information owing to inverse normalization, we carried forward the Pearson residuals to the genome-wide association tests for this trait.

Accounting for complex study design. Additionally, another consideration for model specification in our analysis was accounting for the clustered nature of the data due to the sampling scheme comprising 34 Head Start (HS) programs (primary sampling units) and 280 nested individual centers (sub-clusters). First, a stepwise model selection approach was conducted using HS programs as clusters in a generalized estimating equation (GEE) framework. In this approach, were determined against the inclusion of HS programs as a fixed effect in the mean model. Additionally, we fit two GWA models to determine whether a clustering variable representing the 34 HS programs should be included to account for the complex study design. We used SUGEN to fit the models, which allowed the residual variance in linear regression to differ in different clusters. The first model, which included this variable, resulted in a similar genomic inflation factor (lambda=1.05) as the second model, which did not include the variable (lambda=1.03). Furthermore, the beta coefficients and the standard errors produced by the two models were almost identical. The p-values produced were also relatively similar (Appendix Fig. 13). For these reasons, we decided against accounting for clusters or the sub-clusters in the present analysis.

Genome-wide association study**.** We used three approaches for the genome-wide association (GWA) testing (Fig. 1). Approach 1 (main discovery) was a GWAS in the full study sample. For genetic association testing we used linear and logistic mixed models, respectively for quantitative and binary traits assuming an additive genetic model. We adjusted each model for age, sex, race/ethnicity, first 8 principal components, sugary snacks/beverages, and fluoride content of household water (i.e., fixed effects) (Appendix Table 3). We chose the first 8 principal components for ancestry adjustment, based on evaluation of the scree plot, the parallel coordinates plot, and preliminary association testing results.

Approach 2 (SNP_joint_) investigated single variant associations with ECC testing the hypothesis that a variant has a main and/or interaction effect on ECC using a joint 2-degree-of-freedom (2df) test(Aschard et al. 2010). This approach leveraged potential gene-environment interaction effects in the development of ECC, accounting for the heterogeneity of genetic effects across different strata of interest, i.e. (a) sex, (b) daily between-meal consumption frequency of high versus low sugary snacks and beverages, and (c) exposure to optimal versus sub-optimal level of domestic water source fluoride. The utilization of a joint 2df test evaluated the genetic main effect and the GxE terms jointly (P-joint), primarily testing the null hypothesis (H_0_) of β_G_=β_GxE_=0, which leads to improved power for detecting signals in the context of heterogeneity or GxE interaction, and thus, a higher likelihood of discovering novel genetic variants (Aschard et al. 2010; Sung et al. 2016).

Approach 3 utilized the stratified framework, where the sample was split in two groups, for each of the three environmental exposures listed above. The genetic main effect discovery analysis was performed in each stratum as described in Approach 1.

$$Stratum 1:Y= \beta_{0}^{(1)}+\beta_{G}^{(1)}\times SNP+\beta_{C}^{(1)}\times covariates+e$$

$$Strtatum 2:Y= \beta_{0}^{(2)}+\beta_{G}^{(2)}\times SNP+\beta_{C}^{(2)}\times covariates+e$$

For loci demonstrating genome-wide significant evidence of association in any of the previous analyses, we calculated the p-value for difference (P-difference) between the stratum-specific beta-coefficients of lead SNPs to screen for evidence of GxE interaction effects. The R package EasyStrata was used to perform QC, generate Manhattan and quantile-quantile (Q-Q) plots, and conduct 1-degree-of-freedom (i.e., stratum-specific P-value), 2df (i.e., P-joint), and P-difference tests in the stratified analysis (Winkler et al. 2015).

We used SAIGE for all genetic association analyses because it can account for relatedness using a genetic relationship matrix (i.e., a random effect) and manage unbalanced case-control ratios like the binary clinical decay status trait that has a prevalence of 92%(Zhou et al. 2018). To identify genome-wide significant signals, we excluded variants with MAF<1%, R^2^<0.3 and excluded imputed SNPs with small effective sample sizes (effN<20 for combined and effN<40 for stratified analyses) resulting in the test of ~14 million autosomal SNPs (Appendix Table 4). Assessment of inflation was done using Q-Q plots and genomic inflation (λ_gc_) factors. A multiple testing corrected statistical significance criterion of P<5x10^-8^ was used for all analyses (Uffelmann et al. 2021). Independent genome-wide significant signals within loci were determined based on a linkage disequilibrium (LD) threshold of R^2^<0.6 with the lead SNP. We reported genome-wide significant loci only if the lead SNP had effN≥100 to the reduce likelihood of reporting spurious associations. Finally, we examined the presence of rare variants with functional roles in areas ±250Kb flanking genome-wide significant loci.

The environmental exposures groups are described below with the stratified baseline characteristics presented in Appendix Table 1.

**(a) Sex:** Sex is not a classic environmental factor, but it can be conceptualized as one in genetic epidemiology.

**(b) Daily between-meal consumption frequency of sugary snacks and beverages**: The daily number of sugar-containing snacks and beverages (sugar exposure) consumed by participating children in-between meals was recorded via a questionnaire completed by the parent or legal guardian. We used a dichotomous characterization of this variable with ‘0-1’ category serving as “low sugar”, and the ‘2-3’ and ‘4 or more’ categories as “high sugar”.

**(c) Exposure to optimal level of domestic water source fluoride**: To assess fluoride exposure, we obtained tap water samples from participants’ home water source. The fluoride concentration of the sample was measured by the North Carolina State Laboratory of Public Health using the United States Environment Protection Agency 300.0 method. Only 24% of the participants who underwent clinical examination had data available for fluoride concentration. To impute missing data, we employed a novel machine-learning-based imputation method. The method leveraged geographic information systems (GIS) and combined partitioning around medoids clustering and random forest classification (PAMRF) to impute the remaining data for fluoride concentration of the participants’ water source(Gu et al. 2022). We used fluoride exposure as a dichotomous variable with “0” representing sub-optimal level of fluoride (<0.6ppm) and “1” representing an optimal level (≥0.6ppm) as in a recent investigation (Ha et al. 2019).

**Generalization**. We used summary statistics from two genome-wide meta-analysis of dental caries among predominantly European children and adults for generalization. The child cohort (n=19,000) reported two sets of GWAS summary statistics, namely caries in primary teeth and caries in permanent teeth in children 2.5 to 18 years (Haworth et al. 2018). The adult cohort included approximately 487,000 individuals after combining the multi-study GLIDE consortium association data with UK Biobank data (Shungin et al. 2019). We examined summary estimates of the genome-wide statistically significant SNPs in our study for directional consistency and nominal statistical significance (P<0.05). We considered the variants fulfilling both these criteria as generalized. We did not consider this step as replication and use instead the term “generalization” owing to the differences in the study designs, i.e. the child cohort presented a wider age interval, the adult population in the adult cohort; the primarily European descent sample; and the use of environmental exposures in approaches 2 and 3 in our study in contrast to a conventional discovery GWAS used in the GLIDE studies. Study-specific details can be found in previous reports (Haworth et al. 2018; Shungin et al. 2019). In instances where a lead SNP was not present in the generalization dataset, we identified and interrogated proxy SNPs (R^2^≥0.80 in African, Admixed American and European populations).

**Functional annotation.** We used FUMA GWAS (Functional Mapping and Annotation of Genome-Wide Association Studies) to facilitate functional annotation of single variant testing results (Watanabe et al. 2017). We performed ‘liftover’ of our TOPMed imputed results (build hg38) to hg19 prior to uploading in FUMA. The SNP2GENE process utilized tools including ANNOVAR (Wang et al. 2010), CADD score (Rentzsch et al. 2019), RegulomeDB score (Boyle et al. 2012), and 15-core chromatin state (Ernst and Kellis 2012) to annotate candidate SNPs in risk loci. Additionally, we used FATHMM-XF to evaluate variant pathogenicity (Rogers et al. 2018), HaploReg to inquire further regulatory roles (Ward and Kellis 2016), GTeX to study tissue-specific gene expression (Lonsdale et al. 2013), and GeneCards (Safran et al. 2021) to examine the role of genes in the vicinity of lead signals. We also used the GWAS-catalog (MacArthur et al. 2017) to investigate previously reported associations with lead SNPs or their proxies (R^2^≥0.80).

**Gene-based test and gene-set analysis.** MAGMA v1.6 was used to perform gene-centric analyses within the SNP2GENE process of FUMA. Single variant association testing results were assigned to protein-coding genes obtained from Ensembl build 85 and genome-wide gene-centric significance was set at 0.05/(number of genes tested). The results of this analysis were carried forward to gene-set analyses with gene sets from MsigDB v7.0 for “Curated gene sets” and “GO terms”.

**Pathway enrichment test**. Genes prioritized from SNP2GENE were tested in the GENE2FUNC process using hypergeometric tests to evaluate enrichment in pre-defined gene sets obtained from MsigDB, WikiPathways, and those reported in the GWAS catalog. The protein-coding genes in the FUMA database were used as background and enrichment p-values were adjusted for multiple testing using a False Discovery Rate correction with the Benjamini-Hochberg method.

**Appendix Figures**

**Appendix Figure 1** **|** Flowchart showing enrollment, clinical examinations, and derivation of the GWAS analytical samples in the ZOE 2.0 study, NC, USA. Abbreviations: CIDR: Center for Inherited Disease Research, GWA: Genome-wide Association

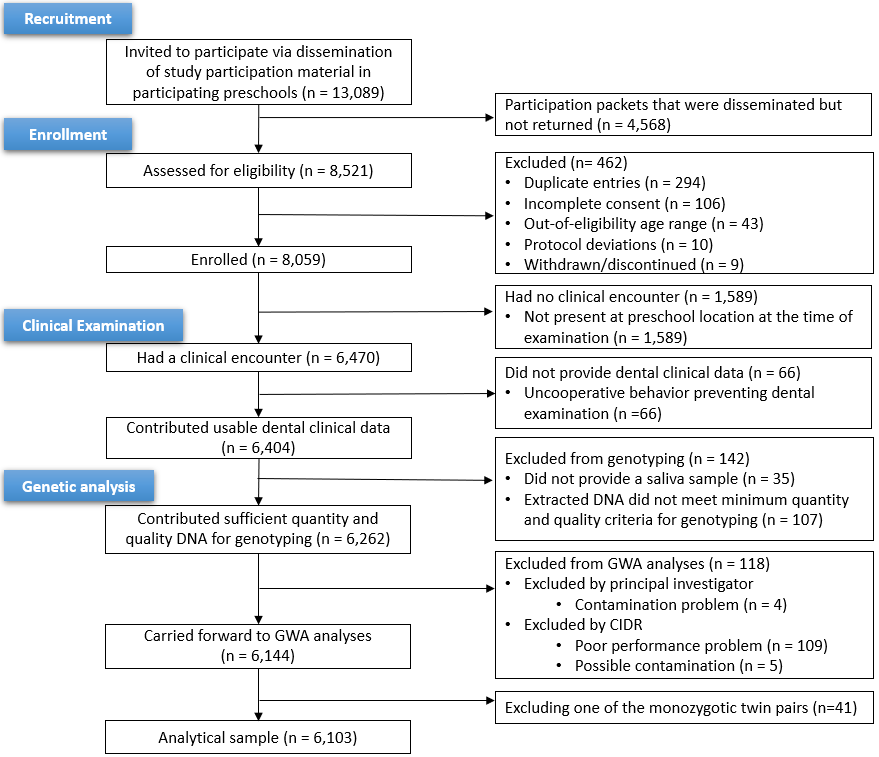

**Appendix Figure 2** **|** Quantile-quantile (Q-Q) plots and the genomic inflation factors (λGC) of the genome-wide association analyses of early childhood caries. (A) Main discovery analysis, (B) Sex-stratified analysis, (C) Fluoride-stratified analysis, (D) Sugar-stratified analysis.

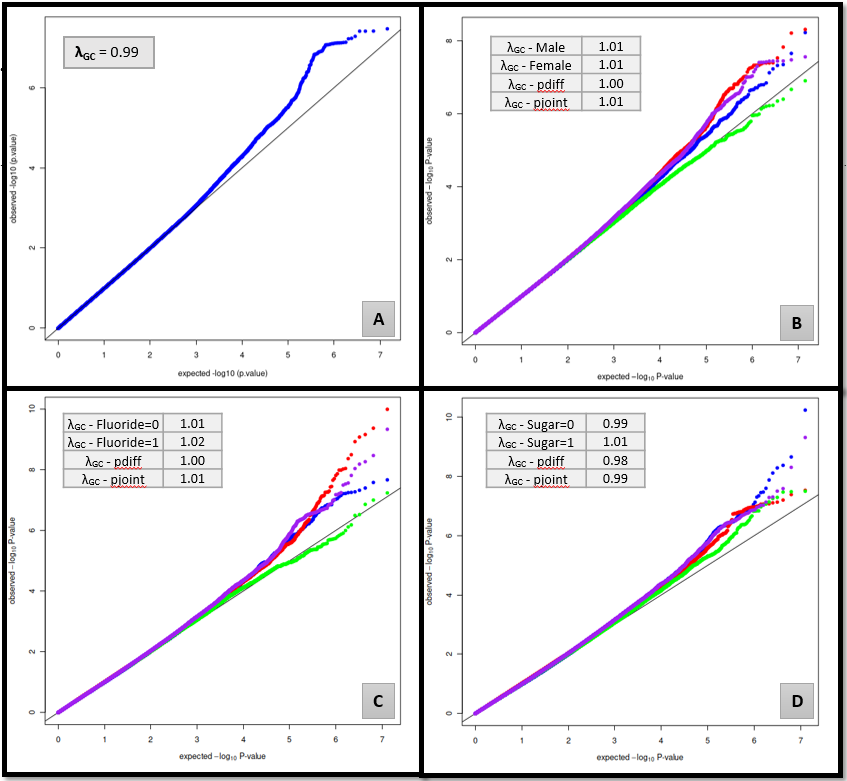

**Appendix Figure 3** **|** Manhattan plots presenting results with genome-wide statistically significant (p<5x10-8) findings for genome-wide association analysis for the “cavitated decay” (d3-6mfs) trait from Approach 1 (Main discovery analysis) and Approach 2 (Joint 2-degree-of-freedom test).The number of significant loci in some plots is more than reported owing to the exclusion of the loci with an effective sample size (effN) less than 100.

| Main discovery analysis | 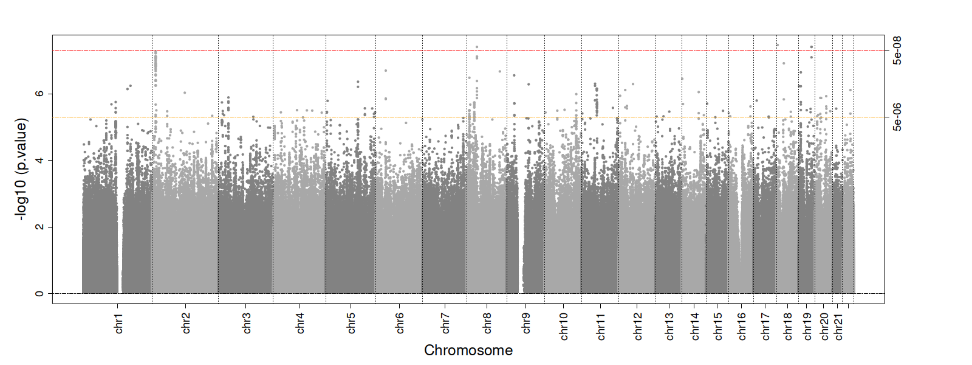 |
| --- | --- |
| Joint test (Sex) | 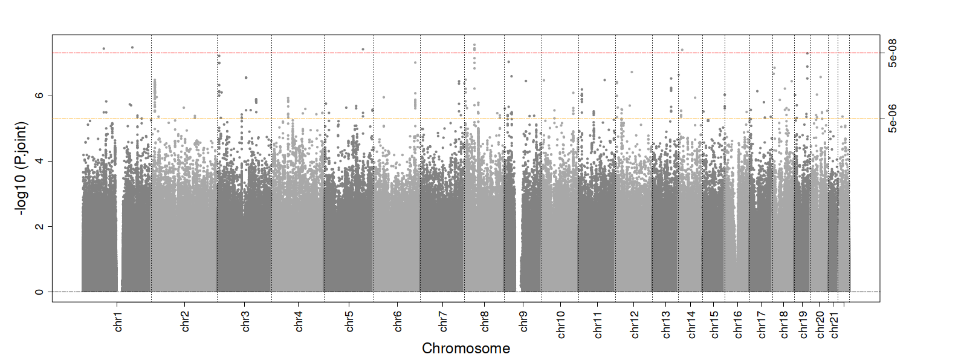 |
| Joint test (Fluoride exposure) | 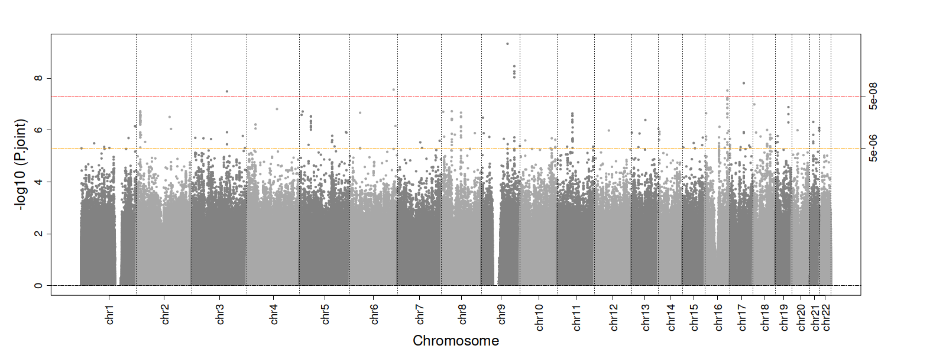 |
| Joint test (Sugar exposure) | 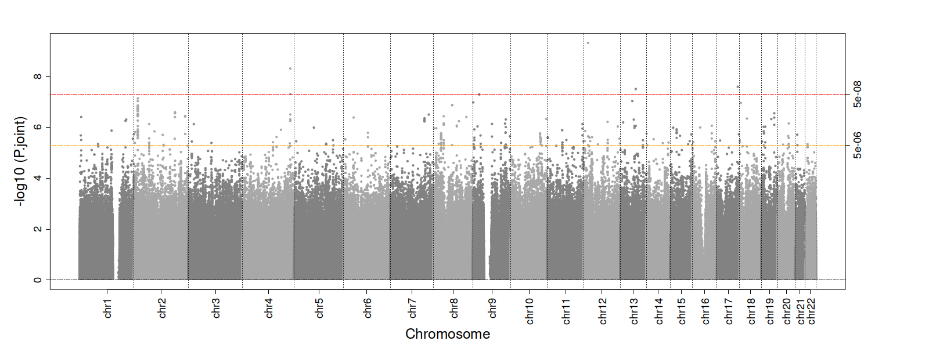 |

**Appendix Figure 4** **|** Manhattan plots presenting results with genome-wide statistically significant (p<5x10-8) findings for genome-wide association analysis for the “cavitated decay” (d3-6mfs) trait in Approach 3 (stratified GWAS). The number of significant loci in some plots is more than reported owing to the exclusion of the loci with an effective sample size (effN) less than 100.

| GWAS-Male | 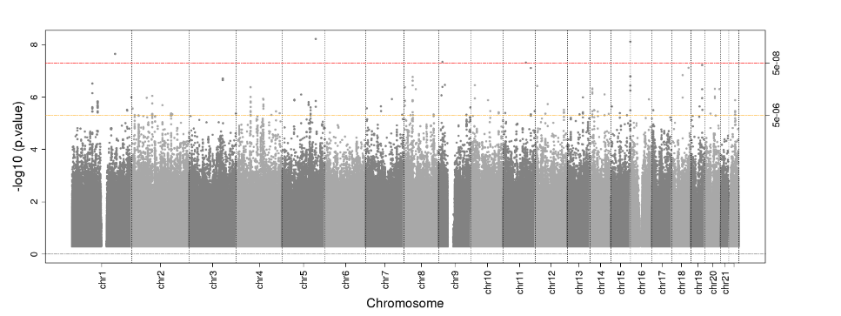 |
| --- | --- |
| GWAS-Female | 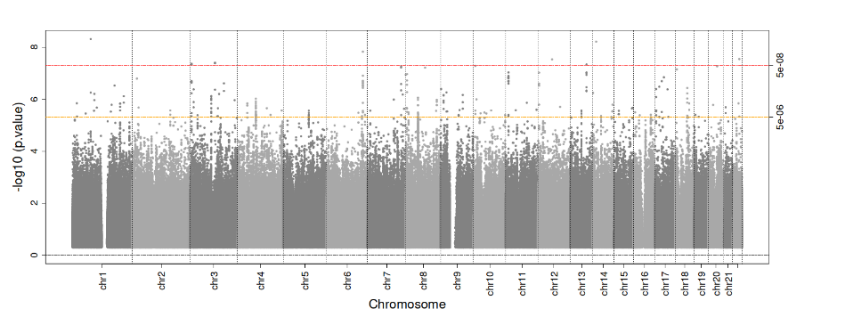 |
| GWAS-Optimal fluoride stratum | 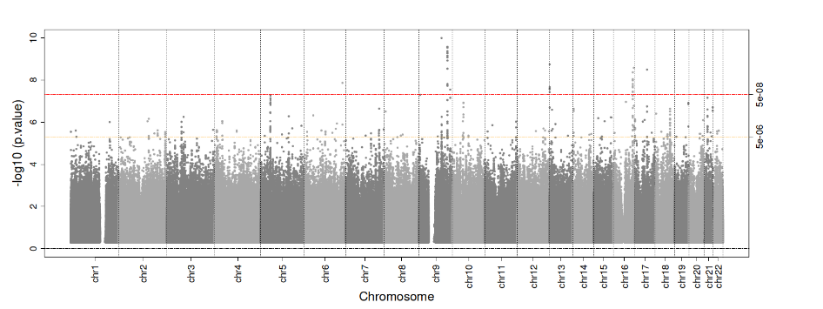 |
| GWAS-Sub-Optimal fluoride stratum | 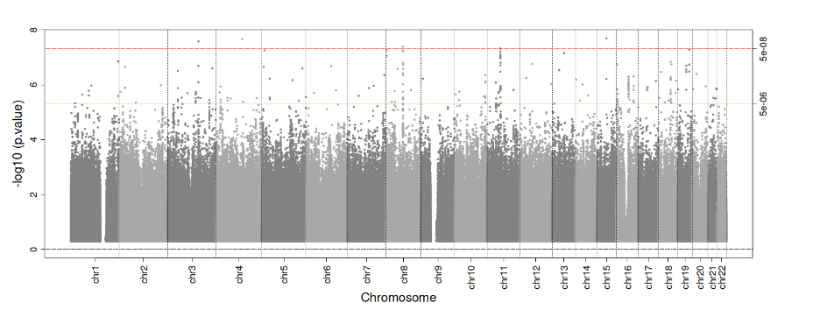 |
| GWAS-High-sugar stratum | 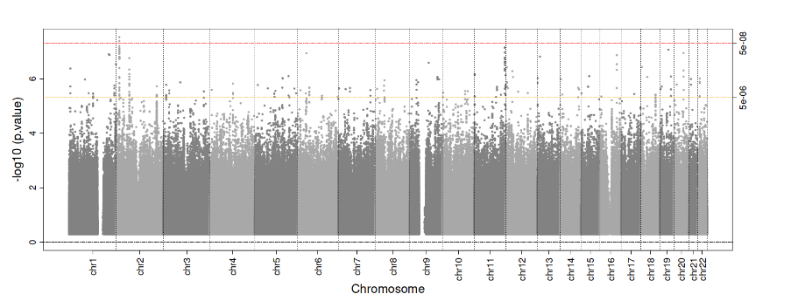 |
| GWAS-Low-sugar stratum | 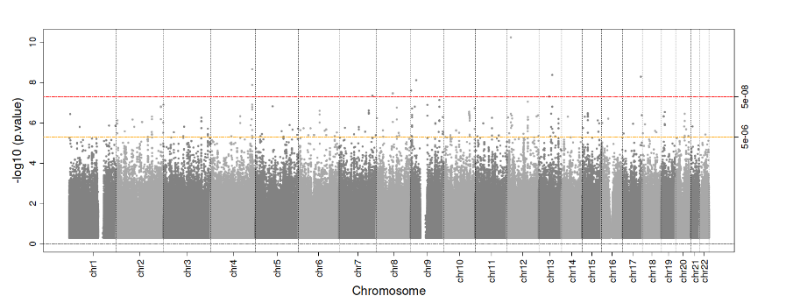 |

**Appendix Figure 5** **|** Results of the gene-centric association analysis using MAGMA (v1.6) illustrating the genome-wide significant association of TAAR6 with the cavitated decay trait among females.

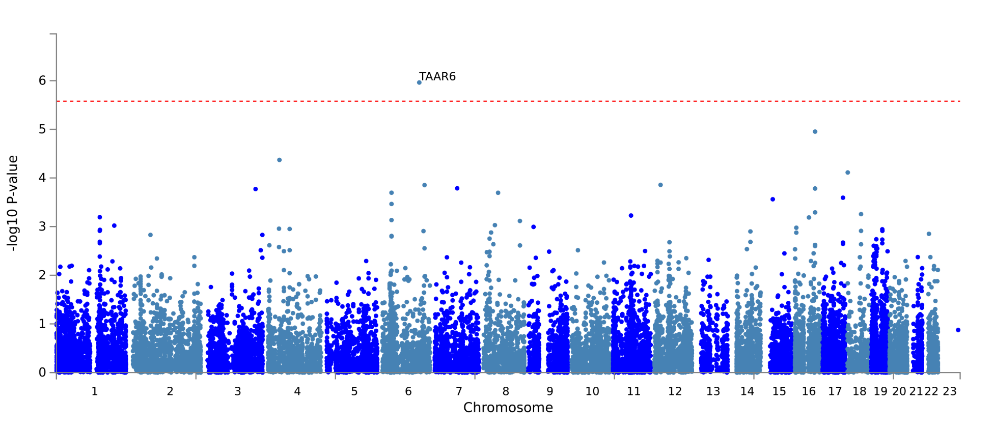

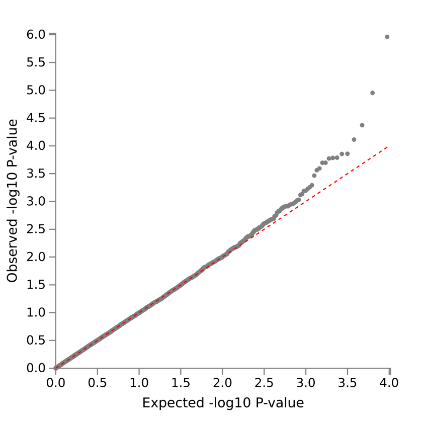

**Appendix Figure 6 |** Identity by descent (IBD) coefficients used to infer participant relatedness. Plot presents kinship coefficients and k_0_ (relationship vector 0) estimates to demonstrate relatedness among related participants. 842 pairs of samples with an estimated φ >1/32 are represented by each point. From top to bottom, the first and second dashed lines encompass the expected full siblings, the second and third lines border the second-degree relative pairs, the third and fourth the expected third-degree relatives and the points under the fourth line are those who are unrelated or related at a fourth or higher degree. Abbreviations: Dup=duplicates, FS=Full siblings, Deg_2_=second degree (half sibling/avuncular/grandparent-grandchild), and Deg_3_=third degree (first cousins), and U=unrelated samples.

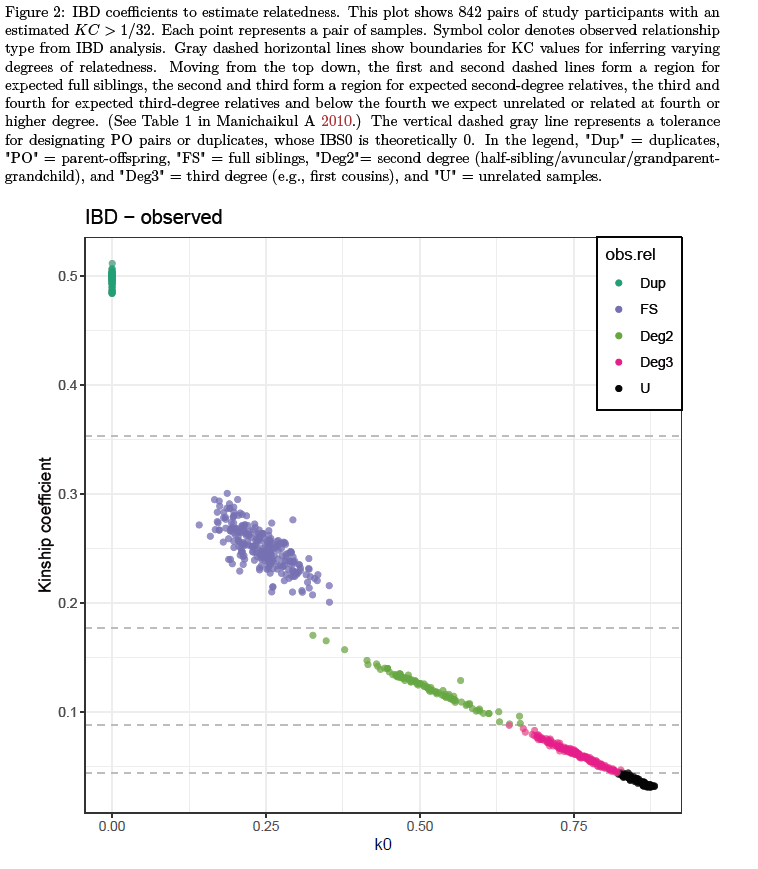

**Appendix Figure 7 |** Scatter plot of the first two eigenvectors from the principal component analysis of 6,144 study participants with 1,204 externally genotyped population controls from HapMap 3 (on the same scale). Left plot: HapMap samples are color-coded according to population; study samples are plotted in light gray. Right plot: Limited to study samples only, color-coded by self-reported race. Axis labels indicate the percentage of variance explained by each eigenvector. (ASW: African ancestry in Southwest USA; CEU: Utah residents with Northern and Western European ancestry from the CEPH collection; CHB: Han Chinese in Beijing, China; CHD: Chinese in Metropolitan Denver, Colorado; GIH: Gujarati Indians in Houston, Texas; JPT: Japanese in Tokyo, Japan; LWK: Luhya in Webuye, Kenya; MXL: Mexican ancestry in Los Angeles, California; MKK: Maasai in Kinyawa, Kenya; TSI: Tuscany in Italy; YRI: Yoruba in Ibadan, Nigeria).

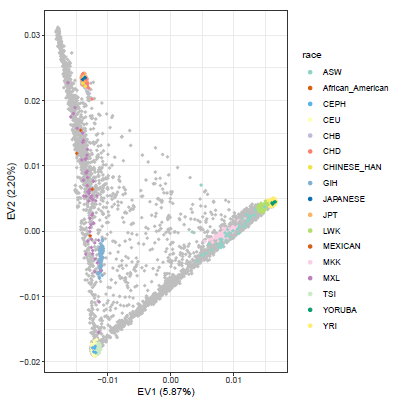

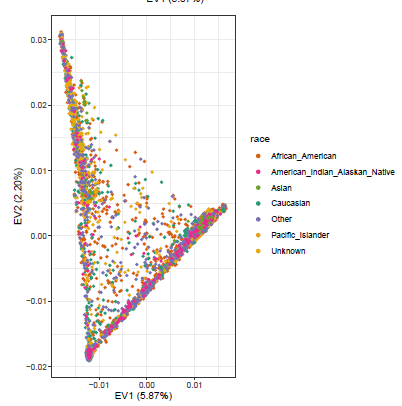

**Appendix Figure 8 |** Estimates of continental-ancestry proportions for the unrelated participants in the ZOE-2.0 study. Each vertical bar is an individual, the four color-coded segments represent the four ancestry populations (European (Blue), Asian (Red), American Admixed (Purple), and African (Green).

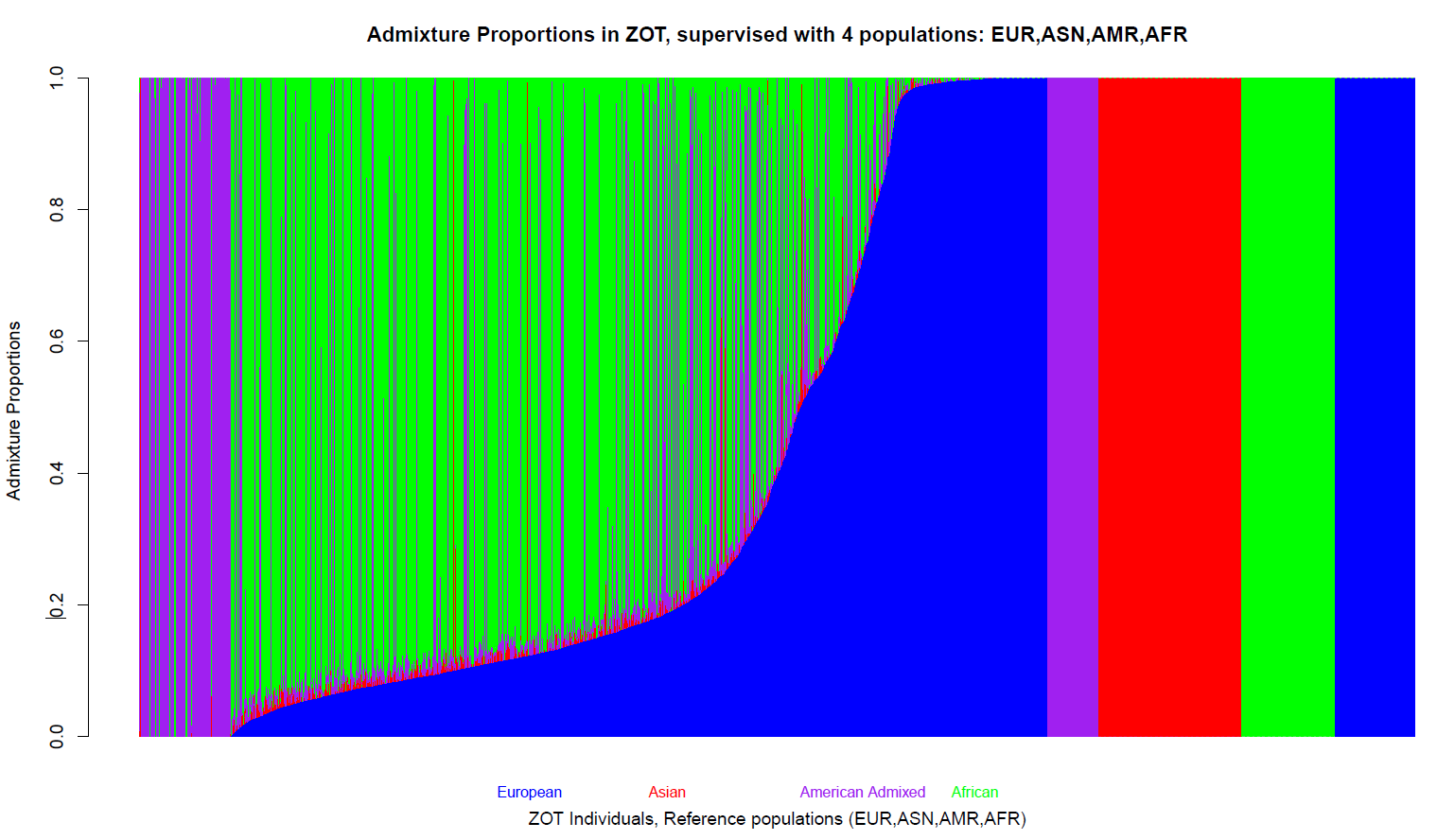

**Appendix Figure 9 |** Flowchart showing the quality control process for the genotyping of the ZOE 2.0 study. Abbreviations: QC: Quality control, QA: Quality assurance, CIDR: Center for Inherited Disease Research, SNP: Single nucleotide polymorphism, MZ: Monozygotic, PCA: Principal component analysis, MCR: Missing call rate, HWE: Hardy-Weinberg equilibrium, MAF: Minor allele frequency

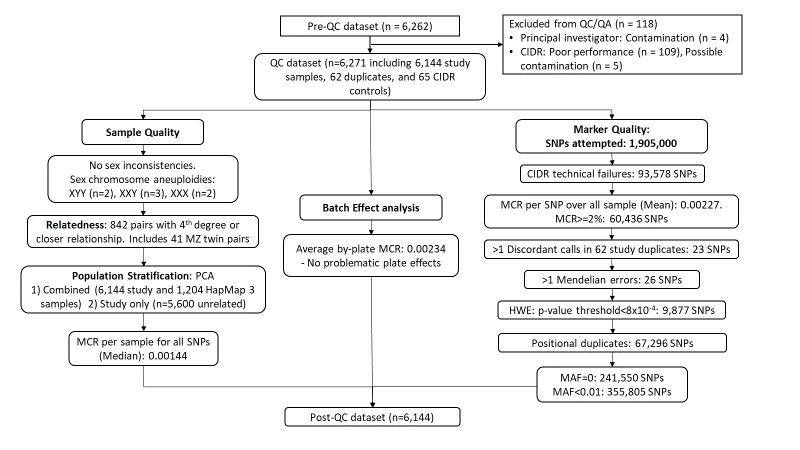

**Appendix Figure 10 |** Distribution of the quantitative clinical decay trait (ICDAS ≥ 1) and cavitated decay trait (ICDAS ≥ 3).

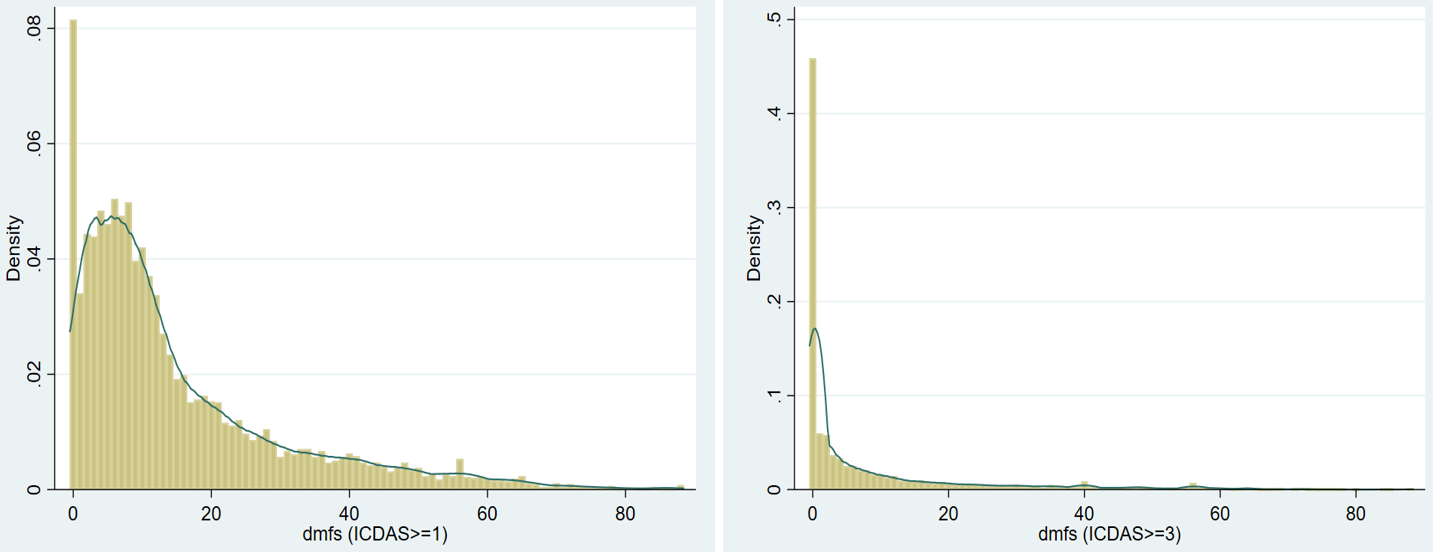

**Appendix Figure 11 |** Diagnostic plots for the linear regression model for clinical decay trait (untransformed) regressed on sex and age in months.

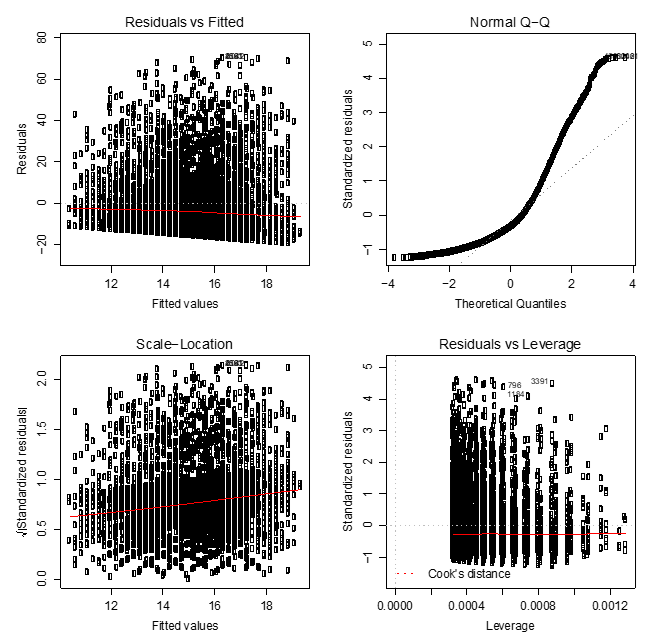

**Appendix Figure 12 |** Diagnostic plots for the linear regression model for clinical decay trait (untransformed) regressed on sex and age in months.

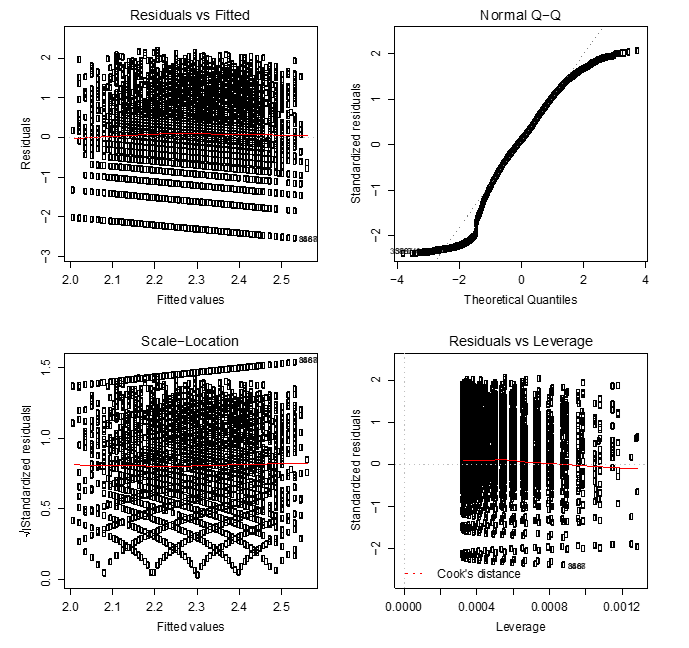

**Appendix Figure 13 |** Comparison of betas (left), standard errors (middle), and p-values (right) obtained from the association tests conducted in the model accounting for the complex study design versus the model not accounting for study design.

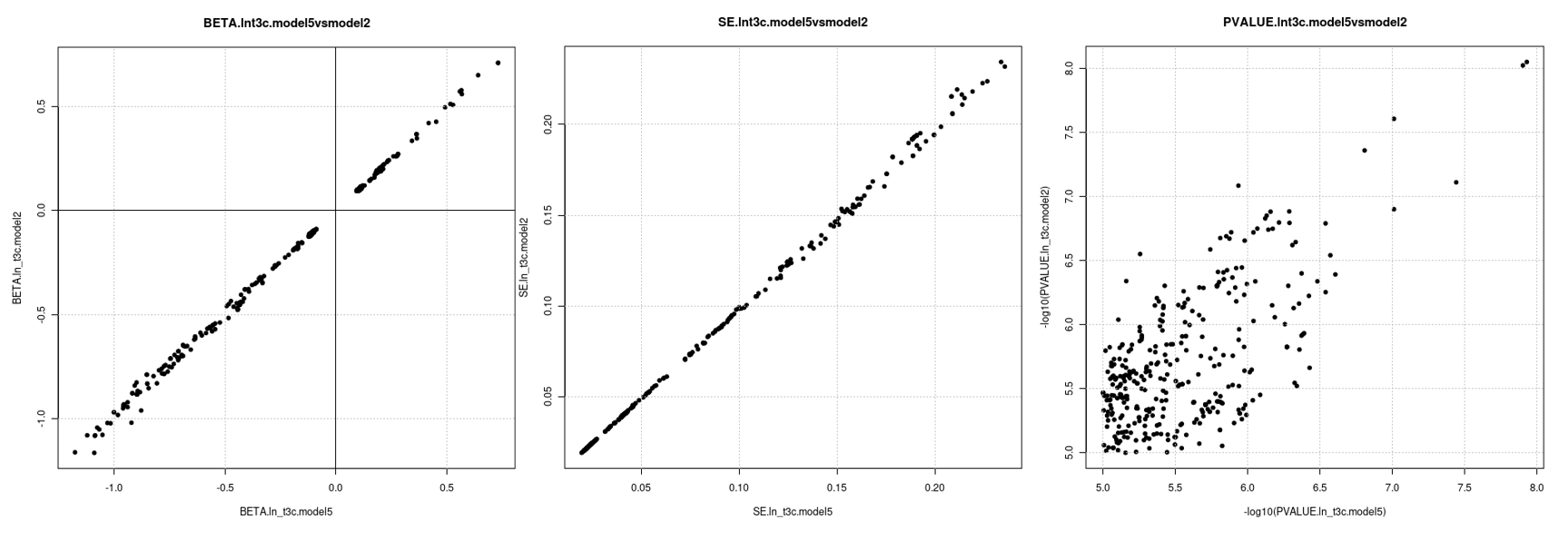

**Appendix Tables**

**Appendix Table 1. Baseline characteristics of the study population (n=6,103), including stratification by sex, fluoride exposure and sugar exposure.**

| **Variable** | **Full sample**  **(n=6,103)** | **Sex-stratified** | | | **Fluoride-stratified** | | | **Sugar-stratified** | |
| --- | --- | --- | --- | --- | --- | --- | --- | --- | --- |
|  |  | **Female (n=3,070)** | **Male (n=3,033)** | **Sub-optimal (n=3,007)** | | **Optimal (n=2,972)** | **Low sugar  (n=1724)** | | **High sugar  (n=4347)** |
| Age (in months), Mean (SD) | 53.5 (7.3) | 53.3 (7.3) | 53.6 (7.2) | 53.7 (7.3) | | 53.3 (7.2) | 53.7 (7.4) | | 53.4 (7.2) |
| Gender, n(%) |  |  |  |  | |  |  | |  |
| Female | 3,070 (50.3) | - | - | 1506 (50.1) | | 1510 (50.8) | 849 (49.3) | | 2182 (50.2) |
| Male | 3,033 (49.7) | - | - | 1501 (49.9) | | 1462 (49.2) | 875 (50.8) | | 2165 (49.8) |
| Race/Ethnicity, n(%)* |  |  |  |  | |  |  | |  |
| Non-Hispanic African American | 2,909 (47.7) | 1,512 (49.3) | 1,397 (46.1) | 1101 (36.6) | | 1757 (59.1) | 695 (40.3) | | 2204 (50.7) |
| Hispanic American | 1,222 (20.0) | 580 (18.9) | 642 (21.2) | 671 (22.3) | | 519 (17.5) | 385 (22.3) | | 827 (19.0) |
| Non- Hispanic White | 1,076 (17.7) | 520 (16.9) | 556 (18.4) | 737 (24.5) | | 325 (10.9) | 385 (22.3) | | 689 (15.9) |
| American Indian/Alaskan Native | 146 (2.4) | 71 (2.3) | 75 (2.5) | 122 (4.1) | | 19 (0.6) | 34 (2.0) | | 111 (2.6) |
| More than one race | 636 (10.4) | 335 (10.9) | 301 (9.9) | 315 (10.5) | | 307 (10.3) | 193 (11.2) | | 442 (10.2) |
| Other | 110 (1.8) | 51 (1.7) | 59 (2.0) | 61 (2.0) | | 45 (1.5) | 32 (1.9) | | 74 (1.7) |
| Fluoride level, n(%) |  |  |  |  | |  |  | |  |
| Sub-optimal (<0.6ppm) | 3,007 (50.3) | 1506 (49.9) | 1501 (50.7) | - | | - | 869 (51.2) | | 2126 (50.0) |
| Optimal (≥0.6ppm) | 2,972 (49.7) | 1510 (50.1) | 1462 (49.3) | - | | - | 829 (48.8) | | 2130 (50.1) |
| Number of between meals sugary snacks and beverage, n(%) |  |  |  |  | |  |  | |  |
| Low sugar (“0-1”) | 1724 (28.4) | 875 (28.6) | 849 (28.2) | 869 (29.0) | | 829 (28.0) | - | | - |
| High sugar (“2 and above”) | 4347 (71.6) | 2182 (71.4) | 2165 (71.8) | 2126 (71.0) | | 2130 (72.0) | - | | - |
| Traits |  |  |  |  | |  |  | |  |
| Clinical decay status, n(%) | 5592 (92.1) | 2807 (91.8) | 2785 (92.3) | 2789 (92.8) | | 2706 (91.1) | 1551 (90.2) | | 4011 (92.8) |
| Cavitated decay status, n(%) | 3281 (54.0) | 1626 (53.2) | 1655 (54.9) | 1732 (57.6) | | 1476 (49.7) | 819 (47.6) | | 2440 (56.4) |
| Clinical decay [d_1-6_mfs, Mean (SD)] | 15.1 (15.4) | 14.4 (14.6) | 15.8 (16.2) | 16.3 (16.3) | | 13.7 (14.2) | 13.4 (14.5) | | 15.7 (15.6) |
| Cavitated decay [d_3-6_mfs, Mean (SD)] | 8.0 (14.2) | 7.6 (13.4) | 8.5 (14.9) | 9.2 (15.4) | | 6.8 (12.7) | 6.7 (13.1) | | 8.5 (14.5) |

Abbreviations. ECC: Early Childhood Caries, SD: Standard deviation; n: number; %: Percentage; ppm: Parts per million; dmfs: decayed, missing, filled surface index

**Appendix Table 2. Overview of the phenotypes along with their prevalence and mean.**

| **Primary Trait** | **Variable type** | **Definition** | **% or Mean (SD)** |
| --- | --- | --- | --- |
| Cavitated decay (mean d_3-6_mfs) | Quantitative (Range: 0-88) | Sum of tooth surface that are decayed (cavitated, ICDAS≥3), missing or filled due to caries. | 8 (14) |
| **Secondary Traits** |  |  |  |
| Clinical decay (mean d_1-6_mfs) | Quantitative (Range: 0-88) | Sum of tooth surfaces that are decayed (non‐cavitated or cavitated lesions, ICDAS ≥1), missing or filled due to caries. | 15 (15) |
| Cavitated decay status (%) | Binary | ≥1 decayed (only cavitated lesions, ICDAS≥3), missing or filled (due to caries) surfaces, in any primary tooth of a child 6 six years. | 54% |
| Clinical decay status (%) | Binary | ≥1 decayed (non‐cavitated or cavitated lesions, ICDAS ≥1), missing or filled (due to caries) surfaces, in any primary tooth of a child under 6 years. | 92% |

**Appendix Table 3: Modelling considerations for quantitative and binary early childhood caries traits.**

| **ECC (caries) traits** | **Range/ Prevalence** | **Distribution** | **Transformation** | **Model** | **Covariates in model** |
| --- | --- | --- | --- | --- | --- |
| Cavitated decay (mean d_3-6_mfs) | 0-88 | zero-inflated, skewed | Pearson residuals* | linear | ev1-ev8, ssbs2, optimalwaterfluorideimputed |
| Clinical decay (mean d_1-6_mfs) | 0-88 | skewed | log transformation | linear | agemo,sex,race,ev1-ev8, ssbs2, optimalwaterfluorideimputed |
| Cavitated decay status (%) | 0.54 | main binary trait | none | logistic | agemo,sex,race,ev1-ev8, ssbs2, optimalwaterfluorideimputed |
| Clinical decay status (%) | 0.92 | “rare” | none | logistic | agemo,sex,race,ev1-ev8, ssbs2, optimalwaterfluorideimputed |

* Generated using zero-inflated negative binomial model and adjusted for age, sex and race/ethnicity.
Abbreviations: ECC: Early childhood caries, ev: Eigenvector (derived from Principal component analysis), ssbs2: Sugary snacks and beverages consumption, optimalwaterfluorideimputed: Fluoride content of household water, agemo: age at examination in months

**Appendix Table 4: Details of genotyping, quality control and statistical analysis**

| **Genotyping** | | | | | | | **Sample quality control and exclusion criteria** | | | | **Prephasing software** | **Imputation** | | | **Filters (No. of SNPs)** |
| --- | --- | --- | --- | --- | --- | --- | --- | --- | --- | --- | --- | --- | --- | --- | --- |
| **DNA source** | **Genotyping array** | **Genotype calling algorithm** | **Inclusion criteria** | | | **Call rate per SNP** | | **Call rate per participant** | **Other** | **SNPs after QC** |  | **SNPs used for imputation** | **Imputation software** | **Reference panel** | All variants (genotyped+imputed): 307,883,017 |
|  |  |  | **MAF** | **Call rate** | **p-value (HWE)** |  |  |  |  |  |  |  |  |  | All non-monomorphic variants (genotyped+imputed): 144,431,368 |
| Saliva | Infinium™ Global Diversity Array-8 v1.0 | GenomeStudio version 2011.1 | **≥**0.01 | **≥**99% | >8e-4 | >99.8% | | >99% | MZ twins | 1,441,911 | Eagle2 V2.4 | 1,142,635 | Minimac4 | TOPMED Reference Panel | MAF>0.01 and R2>0.03 (Main discovery GWAS) 14,371,011 |

Abbreviations: MAF: Minor allele Frequency, HWE: Hardy Weinberg Equilibrium, SNP: Single nucleotide polymorphism, QC: Quality control

**Appendix Table 5: Genomic inflation factor (λ_GC_) for genome-wide analyses performed in this study for Cavitated decay (d_3-6_mfs) trait.**

| **Trait** | **SNPs** | **λ_GC_** |
| --- | --- | --- |
| **Discovery** |  |  |
| Cavitated decay (d_3-6_mfs) | 14371011 | 0.99 |
| **Sex-stratified** |  |  |
| Cavitated decay - Male | 14284873 | 1.01 |
| Cavitated decay - Female | 14439121 | 1.01 |
| Cavitated decay - pdiff | 13998405 | 1.00 |
| Cavitated decay - pjoint | 13998405 | 1.01 |
| **Stratified by:** |  |  |
| **Optimal fluoride in drinking water** |  |  |
| Cavitated decay - Sub-optimal fluoride | 13301965 | 1.00 |
| Cavitated decay - Optimal fluoride | 14321052 | 1.02 |
| Cavitated decay - pdiff | 12919916 | 1.00 |
| Cavitated decay - pjoint | 12919916 | 1.01 |
| **Sugar containing snacks and beverages** |  |  |
| Cavitated decay - Low sugar | 12616116 | 0.99 |
| Cavitated decay - High sugar | 14006303 | 1.01 |
| Cavitated decay - pdiff | 12557725 | 0.98 |
| Cavitated decay - pjoint | 12557725 | 0.99 |

Abbreviations: pdiff: p-value for the difference between the stratum-specific beta coefficients; pjoint: p-value for the joint test

**Appendix Table 6. Novel loci emerging from the genome-wide association analyses conducted for Early Childhood Caries in the ZOE-2.0 study**

| **Trait (Cavitated decay or d_3-6_mfs)** | **Locus** | **rsid** | **SNP (hg38)** | **Chr:Pos (hg19)** | **EA/OA** | **EAF** | **EffN** | **Beta** | **SE** | **p-value** |
| --- | --- | --- | --- | --- | --- | --- | --- | --- | --- | --- |
| **Main discovery (Combined),**  **n=5950** | *DLGAP1* | rs1442369 | chr18:4041807:A:T | 18:4041807 | T/A | 0.40 | 2826 | -0.10 | 0.02 | 3.42E-08 |
|  | *SLC1A5* | rs75906255 | chr19:46796299:G:A | 19:47299556 | A/G | 0.02 | 165 | 0.41 | 0.07 | 3.89E-08 |
|  | *RP11-527N22.1/KCNU1**** | rs58016156 | chr8:37241117:G:A | 8:37098635 | A/G | 0.02 | 312 | 0.31 | 0.06 | 3.91E-08 |
| **Female stratum, n=3002** | *NPAS3*** | rs2255032 | chr14:33119305:A:G | 14:33588511 | G/A | 0.02 | 108 | 0.55 | 0.09 | 6.17E-09 |
|  | *AC016970.1/MIR548A3* | rs79851587 | chr3:103993397:C:A | 3:103712241 | A/C | 0.02 | 133 | 0.46 | 0.08 | 4.04E-08 |
|  | *AC027119.1/MIR4790* | rs76221309 | chr3:5942392:C:T | 3:5984079 | T/C | 0.03 | 198 | 0.38 | 0.07 | 4.35E-08 |
|  | *UBBP5* | rs117286162 | chr13:87201408:T:A | 13:87853663 | A/T | 0.02 | 141 | 0.44 | 0.08 | 4.68E-08 |
| **Male stratum, n=2948** | *AC005740.4*** | rs192232327 | chr5:142064923:C:T | 5:141444488 | T/C | 0.02 | 121 | 0.53 | 0.09 | 5.97E-09 |
|  | *BNC2* | rs191978580 | chr9:16897183:G:A | 9:16897181 | A/G | 0.06 | 303 | 0.34 | 0.06 | 4.49E-08 |
|  | *RP11-856F16.2* | rs74606067 | chr11:94931748:G:T | 11:94664913 | T/G | 0.02 | 118 | 0.50 | 0.09 | 4.75E-08 |
| **Fluoride in water (Sub-optimal stratum), n=2993** | *SLC41A3*** | rs71327750 | chr3:126096213:G:T | 3:125815056 | T/G | 0.14 | 716 | 0.20 | 0.04 | 2.64E-08 |
|  | *NCOA2* | rs13256016 | chr8:70287924:G:A | 8:71200159 | A/G | 0.05 | 296 | 0.31 | 0.06 | 4.04E-08 |
| **Fluoride in water (Optimal stratum), n=2957** | *RP1-62O9.3*** | rs650314 | chr17:49249685:A:C | 17:47327047 | C/A | 0.86 | 691 | -0.22 | 0.04 | 3.22E-09 |
|  | *RP11-525K10.3*** | rs76985043 | chr16:80237146:G:C | 16:80271043 | C/G | 0.02 | 127 | 0.50 | 0.09 | 4.34E-09 |
|  | *ZDHHC14*** | rs3861977 | chr6:157382616:A:T | 6:157803648 | T/A | 0.05 | 292 | 0.33 | 0.06 | 1.37E-08 |
| **Sugary snacks and beverages (≥2) stratum, n = 4252** | *AC092635.1* | rs12052352 | chr2:13537337:G:T | 2:13677462 | T/G | 0.43 | 2065 | 0.12 | 0.02 | 2.95E-08 |

** SNP is also genome-wide statistically significant for the joint test (main genetic effect + gene x environment effect); *** SNP also statistically significant for sex-stratified joint test for this trait; ⴕ Generalization: Nominal significance (p<0.05) + Directional consistency of effect estimates. Abbreviations: ECC:Early Childhood Caries; EA: Effect allele; OA: Other allele; EAF: effect allele frequency; n: sample size; NA: Not applicable; SE: standard error; chr: Chromosome; pos: Position

**Appendix Table 7. Genome-wide statistically significant variants for different analyses for the secondary traits**

| **Trait** | **Locus** | **rsid** | **SNP (hg38)** | **Chr:Pos (hg19)** | **EA/OA** | **EAF** | **effN** | **Beta** | **SE** | **P-value** |
| --- | --- | --- | --- | --- | --- | --- | --- | --- | --- | --- |
| Clinical decay - Female stratum | RNA5SP388/NRXN3 | rs4899701 | chr14:78187579:A:T | 14:78653922 | T/A | 0.31 | 1278 | -0.17 | 0.03 | 3.57E-08 |
| Cavitated decay status – Female stratum | RP11-215I16.1/LOC729506 | rs200747282 | chr5:8173844:C:CA | 5:8173957 | CA/C | 0.51 | 1493 | 0.31 | 0.05 | 1.90E-08 |
| Cavitated decay status – Male stratum | RP11-933H2.4/NUDT16P1 | rs35487488 | chr3:131362117:C:T | 3:131080961 | T/C | 0.09 | 480 | -0.54 | 0.09 | 1.45E-08 |
| Clinical decay – Optimal-fluoride stratum | MACROD1/ RP11-21A7A.2 | rs12420136 | chr11:64043601:C:G | 11:63811073 | G/C | 0.04 | 223 | 0.39 | 0.07 | 2.49E-08 |
| Clinical decay – High-sugar stratum | CTD-2555I5.1** | rs11231965 | chr11:80174062:C:G | 11:79885106 | G/C | 0.11 | 864 | -0.21 | 0.04 | 7.76E-09 |

**SNP is also genome-wide statistically significant for the joint test (main genetic effect + gene x environment effect)

**Appendix Table 8. Novel genetic variants significantly associated with early childhood caries in main discovery GWAS (n=5,950).**

| **Trait** | **Locus** | **Function** | **rsid** | **Chr:Pos (hg19)** | **EA/OA** | **EAF** | **EffN** | **Beta** | **SE** | **P** |
| --- | --- | --- | --- | --- | --- | --- | --- | --- | --- | --- |
| Cavitated decay (d_3-6_mfs) | *DLGAP1* | intronic | rs1442369 | 18:4041807 | T/A | 0.40 | 2826 | -0.10 | 0.02 | 3.42E-08 |
|  | *SLC1A5* | intergenic | rs75906255 | 19:47299556 | A/G | 0.02 | 165 | 0.41 | 0.07 | 3.89E-08 |
|  | *RP11-527N22.1/KCNU1* | intergenic | rs58016156 | 8:37098635 | A/G | 0.02 | 312 | 0.31 | 0.06 | 3.91E-08 |

Abbreviations: ECC:Early Childhood Caries; GWAS: Gwnome-wide Association Study; EA: Effect allele; OA: Other allele; EAF: effect allele frequency; n: sample size; NA: Not applicable; SE: standard error; chr: Chromosome; pos: Position; P: p-value;

**Appendix Table 9: Results of association testing (p-values) for all genome-wide significant SNPs for the primary ECC trait “Cavitated decay” across all traits and analyses.**

| **Locus** | ***RP11-527N22.1**** | ***AC005740.4*** | ***SLC41A3*** | ***ZDHHC14*** | ***RP1-62O9.3*** | ***RP11-525K10.3*** | ***NPAS3*** | ***SLC1A5*** | ***AC092635.1*** | ***DLGAP1*** | ***AC027119.1*** | ***BNC2*** | ***NCOA2*** | ***AC016970.1*** | ***UBBP5*** | ***RP11-856F16.2*** |
| --- | --- | --- | --- | --- | --- | --- | --- | --- | --- | --- | --- | --- | --- | --- | --- | --- |
| **p <5e-8** | 2 | 2 | 2 | 2 | 2 | 2 | 2 | 1 | 1 | 1 | 1 | 1 | 1 | 1 | 1 | 1 |
| **5e-8< p <5e-6** | 4 | 3 | 1 | 1 | 1 | 0 | 0 | 6 | 4 | 4 | 2 | 2 | 1 | 1 | 1 | 1 |
| **5e-6< p <5e-4** | 3 | 6 | 1 | 1 | 0 | 6 | 4 | 2 | 9 | 2 | 2 | 2 | 7 | 5 | 4 | 3 |
| t6c | 3.9E-08 | 2.7E-06 | 9.2E-04 | 5.3E-02 | 1.2E-03 | 6.2E-05 | 3.7E-04 | 3.9E-08 | 5.3E-08 | 3.4E-08 | 1.0E-02 | 2.2E-03 | 3.9E-05 | 1.9E-04 | 1.9E-05 | 1.7E-04 |
| t6 | 1.4E-01 | 2.6E-01 | 3.2E-01 | 2.3E-02 | 4.8E-01 | 9.7E-02 | 1.2E-01 | 3.1E-02 | 4.0E-02 | 1.3E-02 | 2.7E-01 | 1.6E-02 | 8.7E-01 | 4.1E-01 | 6.6E-01 | 6.0E-01 |
| t3c | 5.6E-02 | 1.2E-03 | 2.7E-01 | 5.3E-02 | 3.3E-01 | 1.1E-02 | 1.9E-02 | 7.8E-03 | 8.9E-05 | 1.5E-02 | 1.5E-01 | 2.8E-02 | 6.5E-03 | 4.3E-02 | 8.3E-02 | 2.1E-01 |
| t3 | 4.5E-01 | 5.1E-01 | 5.6E-01 | 9.6E-01 | 5.9E-01 | 4.6E-01 | 7.5E-02 | 4.6E-01 | 3.2E-02 | 8.1E-01 | 7.4E-01 | 5.8E-01 | 1.2E-01 | 7.6E-01 | 7.6E-01 | 9.0E-01 |
| t6c_SS_male | 3.8E-06 | 6.0E-09 | 1.6E-03 | 5.3E-02 | 5.7E-03 | 4.5E-02 | 5.7E-01 | 5.9E-08 | 2.1E-04 | 2.7E-04 | 7.1E-02 | 4.5E-08 | 1.9E-03 | 8.9E-01 | 6.4E-01 | 4.7E-08 |
| t6c_SS_female | 2.5E-04 | 5.7E-01 | 1.6E-01 | 3.3E-01 | 5.4E-02 | 1.9E-04 | 6.2E-09 | 4.0E-02 | 6.0E-05 | 3.0E-05 | 4.3E-08 | 1.2E-01 | 1.5E-02 | 4.0E-08 | 4.7E-08 | 8.8E-01 |
| t6c_SS_P_diff | 4.1E-01 | 1.8E-04 | 2.0E-01 | 5.1E-01 | 5.3E-01 | 2.5E-01 | 5.4E-06 | 2.5E-02 | 8.6E-01 | 6.9E-01 | 4.0E-07 | 7.4E-07 | 7.0E-01 | 1.5E-04 | 5.7E-04 | 3.7E-05 |
| t6c_SS_P_joint | 2.8E-08 | 3.8E-08 | 2.6E-03 | 9.5E-02 | 3.4E-03 | 1.2E-04 | 3.9E-08 | 5.0E-08 | 3.3E-07 | 2.1E-07 | 6.1E-08 | 9.2E-08 | 4.2E-04 | 2.8E-07 | 3.0E-07 | 3.3E-07 |
| t6_SS_male | 2.0E-01 | 9.8E-02 | 1.4E-01 | 5.7E-02 | 4.8E-01 | 6.3E-01 | 7.6E-01 | 1.4E-03 | 5.9E-01 | 8.7E-02 | 5.2E-01 | 2.4E-04 | 9.3E-01 | 4.8E-01 | 1.1E-01 | 1.2E-01 |
| t6_SS_female | 3.8E-01 | 8.9E-01 | 9.9E-01 | 1.5E-01 | 7.5E-01 | 6.0E-02 | 1.0E-02 | 8.1E-01 | 1.7E-02 | 6.2E-02 | 3.2E-02 | 6.6E-01 | 6.6E-01 | 9.9E-02 | 3.1E-02 | 4.8E-01 |
| t6_SS_P_diff | 7.6E-01 | 2.7E-01 | 3.0E-01 | 7.8E-01 | 7.9E-01 | 3.3E-01 | 3.9E-02 | 1.8E-02 | 1.9E-01 | 8.9E-01 | 5.1E-02 | 4.0E-03 | 7.0E-01 | 9.7E-02 | 7.4E-03 | 1.1E-01 |
| t6_SS_P_joint | 3.0E-01 | 2.5E-01 | 3.4E-01 | 5.7E-02 | 7.4E-01 | 1.5E-01 | 3.6E-02 | 5.9E-03 | 5.0E-02 | 4.1E-02 | 8.1E-02 | 1.1E-03 | 9.1E-01 | 2.0E-01 | 2.7E-02 | 2.4E-01 |
| t3c_SS_male | 2.3E-01 | 1.2E-05 | 2.8E-02 | 2.9E-02 | 7.2E-01 | 2.1E-01 | 7.3E-01 | 7.2E-05 | 7.3E-03 | 1.8E-02 | 1.2E-01 | 3.5E-04 | 1.2E-01 | 6.4E-01 | 8.2E-01 | 3.6E-03 |
| t3c_SS_female | 9.6E-02 | 7.7E-01 | 6.3E-01 | 5.4E-01 | 2.4E-01 | 2.1E-02 | 2.0E-04 | 6.2E-01 | 3.8E-03 | 2.0E-01 | 3.9E-04 | 7.3E-01 | 3.4E-02 | 1.6E-03 | 9.6E-03 | 3.1E-01 |
| t3c_SS_P_diff | 7.8E-01 | 3.3E-03 | 5.3E-02 | 2.6E-01 | 5.7E-01 | 4.9E-01 | 3.8E-03 | 1.7E-03 | 9.2E-01 | 4.4E-01 | 3.7E-04 | 6.0E-03 | 6.5E-01 | 1.3E-02 | 5.0E-02 | 4.2E-03 |
| t3c_SS_P_joint | 1.2E-01 | 6.4E-05 | 7.9E-02 | 7.6E-02 | 4.7E-01 | 3.3E-02 | 9.4E-04 | 3.4E-04 | 4.2E-04 | 2.6E-02 | 5.7E-04 | 1.6E-03 | 3.2E-02 | 6.1E-03 | 3.4E-02 | 8.5E-03 |
| t3_SS_male | 9.5E-01 | 2.0E-01 | 8.7E-01 | 7.6E-01 | 3.7E-01 | 3.2E-01 | 4.3E-01 | 9.1E-01 | 1.2E-01 | 8.9E-01 | 5.3E-01 | 6.8E-01 | 1.6E-01 | 9.9E-02 | 5.2E-01 | 4.7E-01 |
| t3_SS_female | 2.8E-01 | 7.1E-01 | 4.4E-01 | 7.7E-01 | 8.5E-01 | 8.7E-01 | 8.0E-02 | 1.9E-01 | 1.5E-01 | 8.7E-01 | 2.9E-01 | 2.5E-01 | 5.4E-01 | 6.5E-02 | 8.6E-01 | 3.9E-01 |
| t3_SS_P_diff | 4.2E-01 | 2.4E-01 | 6.7E-01 | 6.7E-01 | 4.4E-01 | 5.4E-01 | 5.0E-01 | 3.1E-01 | 9.1E-01 | 9.9E-01 | 2.4E-01 | 2.7E-01 | 5.9E-01 | 1.4E-02 | 5.5E-01 | 2.6E-01 |
| t3_SS_P_joint | 5.6E-01 | 4.1E-01 | 7.3E-01 | 9.1E-01 | 6.6E-01 | 6.0E-01 | 1.6E-01 | 4.2E-01 | 1.1E-01 | 9.8E-01 | 4.7E-01 | 4.7E-01 | 3.1E-01 | 4.7E-02 | 8.0E-01 | 5.3E-01 |
| t6c_ssbs2_str_ssbs20 | 1.5E-02 | 3.3E-01 | 5.4E-01 | 2.8E-01 | 8.0E-02 | 2.7E-02 | 3.9E-01 | 1.6E-02 | 1.5E-01 | 1.3E-02 | 2.5E-02 | 8.1E-01 | 4.6E-01 | 4.1E-03 | 2.0E-03 | 1.4E-01 |
| t6c_ssbs2_str_ssbs21 | 1.1E-06 | 8.0E-07 | 2.6E-04 | 1.5E-01 | 5.2E-03 | 1.0E-03 | 2.1E-04 | 8.2E-07 | 3.0E-08 | 3.7E-07 | 6.7E-02 | 8.5E-04 | 1.4E-05 | 5.9E-03 | 1.0E-03 | 4.8E-04 |
| t6c_ssbs2_str_P_diff | 7.0E-01 | 3.5E-02 | 1.4E-01 | 9.9E-01 | 9.9E-01 | 9.8E-01 | 1.3E-01 | 2.8E-01 | 7.0E-02 | 5.5E-01 | 3.5E-01 | 1.0E-01 | 5.5E-02 | 4.6E-01 | 6.1E-01 | 4.9E-01 |
| t6c_ssbs2_str_P_joint | 3.7E-07 | 3.2E-06 | 1.0E-03 | 2.0E-01 | 4.4E-03 | 3.9E-04 | 7.1E-04 | 2.8E-07 | 7.4E-08 | 1.1E-07 | 1.5E-02 | 3.7E-03 | 6.2E-05 | 3.6E-04 | 4.0E-05 | 7.7E-04 |
| t6_ssbs2_str_ssbs20 | . | . | 2.0E-01 | 4.2E-02 | 5.3E-01 | . | . | . | 3.7E-01 | 5.8E-01 | . | 2.6E-01 | 3.8E-01 | . | 2.0E-01 | . |
| t6_ssbs2_str_ssbs21 | . | . | 7.2E-01 | 1.9E-01 | 6.0E-01 | . | . | . | 5.0E-02 | 9.0E-03 | . | 2.5E-02 | 7.4E-01 | . | 1.2E-01 | . |
| t6_ssbs2_str_P_diff | . | . | 3.7E-01 | 3.8E-01 | 7.9E-01 | . | . | . | 7.6E-01 | 3.6E-01 | . | 7.6E-01 | 3.6E-01 | . | 4.8E-02 | . |
| t6_ssbs2_str_P_joint | . | . | 4.2E-01 | 5.4E-02 | 7.1E-01 | . | . | . | 9.8E-02 | 2.8E-02 | . | 4.3E-02 | 6.5E-01 | . | 1.3E-01 | . |
| t3c_ssbs2_str_ssbs20 | 8.8E-02 | 7.6E-01 | 7.9E-01 | 6.6E-02 | 5.4E-01 | 6.4E-01 | 5.1E-01 | 9.7E-01 | 1.1E-01 | 4.8E-01 | 5.7E-01 | 7.4E-01 | 9.9E-01 | 1.5E-01 | 5.9E-01 | 3.2E-01 |
| t3c_ssbs2_str_ssbs21 | 2.6E-01 | 1.2E-04 | 1.2E-01 | 3.5E-01 | 3.9E-01 | 5.9E-03 | 1.2E-02 | 1.1E-03 | 3.3E-04 | 1.4E-02 | 3.0E-02 | 2.1E-02 | 7.8E-04 | 1.4E-01 | 8.4E-02 | 3.7E-02 |
| t3c_ssbs2_str_P_diff | 3.5E-01 | 5.0E-02 | 2.9E-01 | 3.4E-01 | 9.3E-01 | 2.7E-01 | 3.6E-01 | 5.4E-02 | 5.7E-01 | 5.0E-01 | 1.0E-01 | 3.3E-01 | 5.4E-02 | 7.2E-01 | 5.6E-01 | 4.9E-02 |
| t3c_ssbs2_str_P_joint | 1.2E-01 | 5.9E-04 | 2.9E-01 | 1.2E-01 | 5.7E-01 | 2.0E-02 | 3.4E-02 | 4.9E-03 | 4.4E-04 | 3.7E-02 | 8.1E-02 | 6.5E-02 | 3.6E-03 | 1.2E-01 | 2.0E-01 | 6.9E-02 |
| t3_ssbs2_str_ssbs20 | . | . | . | . | . | . | . | . | 2.3E-01 | 5.1E-01 | . | . | . | . | . | . |
| t3_ssbs2_str_ssbs21 | . | . | . | . | . | . | . | . | 7.2E-02 | 4.9E-01 | . | . | . | . | . | . |
| t3_ssbs2_str_P_diff | . | . | . | . | . | . | . | . | 9.2E-01 | 3.5E-01 | . | . | . | . | . | . |
| t3_ssbs2_str_P_joint | . | . | . | . | . | . | . | . | 9.6E-02 | 6.3E-01 | . | . | . | . | . | . |
| t6c_fluo_str_fluo0 | 1.2E-04 | 6.3E-05 | 2.6E-08 | 1.1E-01 | 3.4E-01 | 6.6E-01 | 8.3E-03 | 5.4E-08 | 4.4E-05 | 3.0E-06 | 1.2E-01 | 7.7E-03 | 4.0E-08 | 7.4E-02 | 4.8E-04 | 1.3E-03 |
| t6c_fluo_str_fluo1 | 5.5E-05 | 5.1E-02 | 6.0E-02 | 1.4E-08 | 3.2E-09 | 4.3E-09 | 8.2E-03 | 1.4E-01 | 1.6E-04 | 1.3E-03 | 3.4E-02 | 2.2E-01 | 4.4E-01 | 1.0E-04 | 6.2E-03 | 4.9E-02 |
| t6c_fluo_str_P_diff | 6.4E-01 | 3.9E-01 | 3.2E-07 | 5.8E-08 | 2.9E-06 | 3.4E-05 | 9.0E-01 | 4.0E-02 | 9.0E-01 | 3.4E-01 | 7.2E-01 | 3.7E-01 | 7.5E-05 | 8.6E-02 | 9.8E-01 | 5.7E-01 |
| t6c_fluo_str_P_joint | 1.9E-07 | 4.9E-05 | 3.2E-08 | 2.8E-08 | 1.5E-08 | 3.0E-08 | 9.4E-04 | 1.3E-07 | 1.9E-07 | 1.0E-07 | 3.2E-02 | 1.3E-02 | 2.1E-07 | 1.1E-04 | 5.3E-05 | 8.3E-04 |
| t6_fluo_str_fluo0 | 2.0E-01 | 4.2E-01 | 4.3E-02 | 9.1E-01 | 2.3E-01 | 8.1E-01 | 5.3E-01 | . | 6.3E-02 | 4.1E-02 | 9.1E-01 | 3.8E-02 | 7.0E-01 | 8.3E-01 | 4.5E-01 | . |
| t6_fluo_str_fluo1 | 3.6E-01 | 3.6E-01 | 4.8E-01 | 6.1E-04 | 3.9E-02 | 3.8E-03 | 9.4E-02 | . | 2.4E-01 | 1.1E-01 | 9.1E-02 | 1.4E-01 | 4.2E-01 | 3.1E-01 | 8.2E-02 | . |
| t6_fluo_str_P_diff | 6.7E-01 | 8.3E-01 | 5.7E-02 | 1.0E-02 | 2.2E-02 | 1.9E-02 | 4.3E-01 | . | 6.4E-01 | 7.6E-01 | 2.1E-01 | 7.0E-01 | 3.8E-01 | 5.3E-01 | 6.6E-02 | . |
| t6_fluo_str_P_joint | 2.9E-01 | 4.7E-01 | 1.0E-01 | 2.8E-03 | 5.7E-02 | 1.5E-02 | 2.0E-01 | . | 8.9E-02 | 3.5E-02 | 2.4E-01 | 4.0E-02 | 6.7E-01 | 5.8E-01 | 1.7E-01 | . |
| t3c_fluo_str_fluo0 | 1.4E-01 | 3.0E-03 | 2.5E-02 | 5.9E-01 | 1.4E-01 | 1.0E+00 | 7.3E-02 | 1.9E-02 | 2.8E-03 | 1.3E-02 | 6.8E-01 | 5.0E-02 | 3.8E-05 | 6.8E-01 | 4.3E-01 | 7.6E-01 |
| t3c_fluo_str_fluo1 | 2.1E-01 | 1.5E-01 | 3.1E-01 | 2.0E-04 | 5.5E-03 | 1.1E-04 | 1.1E-01 | 2.2E-01 | 6.5E-03 | 3.3E-01 | 1.3E-01 | 2.3E-01 | 3.6E-01 | 7.2E-03 | 5.0E-02 | 9.7E-02 |
| t3c_fluo_str_P_diff | 7.0E-01 | 5.0E-01 | 2.4E-02 | 1.3E-03 | 3.3E-03 | 3.7E-03 | 9.2E-01 | 6.6E-01 | 8.7E-01 | 2.9E-01 | 4.6E-01 | 6.3E-01 | 1.1E-03 | 8.8E-02 | 3.0E-01 | 2.9E-01 |
| t3c_fluo_str_P_joint | 1.5E-01 | 4.3E-03 | 4.8E-02 | 8.4E-04 | 7.3E-03 | 5.6E-04 | 5.7E-02 | 3.1E-02 | 2.8E-04 | 2.9E-02 | 3.0E-01 | 7.2E-02 | 1.4E-04 | 2.5E-02 | 1.1E-01 | 2.4E-01 |
| t3_fluo_str_fluo0 | 5.8E-01 | 9.8E-01 | 7.9E-01 | 6.2E-01 | 2.9E-01 | 6.3E-01 | 2.7E-02 | 6.5E-01 | 5.1E-02 | 8.5E-01 | 6.9E-01 | 4.8E-01 | 8.9E-02 | 8.7E-01 | 8.1E-01 | 1.6E-01 |
| t3_fluo_str_fluo1 | 5.6E-01 | 3.6E-01 | 5.3E-01 | 6.2E-01 | 8.9E-01 | 6.3E-01 | 7.8E-01 | 6.0E-01 | 2.7E-01 | 6.6E-01 | 4.4E-01 | 9.4E-01 | 7.1E-01 | 4.1E-01 | 8.8E-01 | 1.3E-01 |
| t3_fluo_str_P_diff | 9.0E-01 | 4.9E-01 | 7.8E-01 | 4.8E-01 | 3.7E-01 | 9.8E-01 | 1.7E-01 | 8.8E-01 | 5.1E-01 | 6.6E-01 | 4.2E-01 | 5.6E-01 | 4.3E-01 | 4.7E-01 | 9.7E-01 | 4.0E-02 |
| t3_fluo_str_P_joint | 7.3E-01 | 6.6E-01 | 8.0E-01 | 7.8E-01 | 5.7E-01 | 8.0E-01 | 8.3E-02 | 7.9E-01 | 8.1E-02 | 8.9E-01 | 6.9E-01 | 7.7E-01 | 2.2E-01 | 7.0E-01 | 9.6E-01 | 1.2E-01 |

p: p-value for single variant association testing; *Genome-wide significant in Approaches 1 and 2; Traits - t6c: Cavitated decay,,t3c: Clinical decay,,t6: Cavitated decay status (Yes/No), t3: Clinical decay status (Yes/No); Environmental factors - SS: Sex-stratified analysis, ssbs2_str: Sugary snacks and beverages in-between meals (Sugar-stratified analysis), fluo_str: Fluoride level in water (Fluoride stratified analysis), ssbs20 - Low sugar stratum, ssbs21 - High sugar stratum, fluo0 - Sub-optimal fluoride stratum, fluo1 - Optimal fluoride stratum; P_joint: p-value for joint (2df) test (testing Genetic + Gene-environment interaction effect jointly), P_diff: p-value for difference in betas, testing interaction effect

**Appendix Table 10: Generalization of loci from ZOE-2.0 main discovery GWAS (Approach 1) to GLIDE children and GLIDE adults.**

| **ZOE-2.0 study (N=5950), Trait: Cavitated decay** | | | | | | | **GLIDE children (Caries yes/no trait - Primary teeth)** | | | | **GLIDE children (Caries yes/no trait - Permanent teeth)** | | | | | **GLIDE adults (DMFS and Dentures traits)** | | | |
| --- | --- | --- | --- | --- | --- | --- | --- | --- | --- | --- | --- | --- | --- | --- | --- | --- | --- | --- | --- |
| Locus | rsid | EA/OA | EAF | Beta | P | Beta | | P | N | DC | Beta | P | N | DC | Beta | | P | N | DC |
| *DLGAP1* | rs1442369 | T/A | 0.40 | -0.10 | 3.42E-08 | -0.02 | | 0.62 | 13920 | No | 0.01 | 0.78 | 13172 | Yes | 0.01 | | **0.03** | 285246 | No |
| *SLC1A5* | rs75906255 | A/G | 0.02 | 0.41 | 3.89E-08 | -0.07 | | 0.39 | 17921 | No | -0.10 | 0.22 | 13365 | No | -0.00 | | 0.94 | 285248 | No |
| *RP11-527N22.1/*  *KCNU1* | rs58016156 | A/G | 0.02 | 0.31 | 3.91E-08 | -0.52 | | 0.34 | 1923 | No | * | * | * | * | * | | * | * | * |

*Proxy SNP with R^2^>0.8 not found; **SNP is also genome-wide statistically significant for the joint test (main genetic effect + gene x environment effect).
Generalization: Nominal Significance + Directional consistency, GLIDE children study: Caries Yes/No trait.
Abbreviations: DMFS: Decayed, Missing, and Filled Surfaces; EA: Effect allele; OA: Other allele; EAF: effect allele frequency; N: sample size; NA: Not applicable; SE: standard error; chr: Chromosome; pos: Position; DC: Directional consistency

**Appendix Table 11: Generalization of loci from ZOE-2.0 GWAS accounting for sex (Approach 2 and 3) to GLIDE children and GLIDE adults.**

|  | | | | | | **GLIDE children (Primary teeth)** | | | | **GLIDE children (Permanent teeth)** | | | | **GLIDE adults (DMFS and Dentures traits)** | | | |
| --- | --- | --- | --- | --- | --- | --- | --- | --- | --- | --- | --- | --- | --- | --- | --- | --- | --- |
| **Locus** | **rsid** | **EA/OA** | **EAF** | **Beta** | **P** | **Beta** | **P** | **N** | **DC** | **Beta** | **P** | **N** | **DC** | **Beta** | **P** | **N** | **DC** |
| **Trait (analysis): Cavitated decay (GWAS - Male stratum)** | | | | | | | | | | | | | | | | | |
| *AC005740.4* | rs192232327 | T/C | 0.02 | 0.53 | 5.97E-09 | - | - | - | - | - | - | - | - | - | - | - | - |
| *BNC2* | rs191978580 | A/G | 0.06 | 0.34 | 4.49E-08 | 0.07 | 0.71 | 4867 | Yes | 0.26 | 0.57 | 822 | No | 0.01 | 0.24 | 285249 | No |
| *RP11-856F16.2* | rs74606067 | T/G | 0.02 | 0.5 | 4.75E-08 | 0.14 | 0.08 | 13920 | Yes | 0.02 | 0.73 | 13344 | Yes | 0.01 | **0.01** | 285248 | Yes |
| **Cavitated decay (GWAS - Female stratum)** | | | | | | | | | | | | | | | | | |
| *NPAS3*** | rs2255032 | G/A | 0.02 | 0.55 | 6.17E-09 | -0.09 | 0.27 | 18998 | Yes | -0.09 | 0.33 | 13385 | Yes | 0 | 0.66 | 285248 | No |
| *AC016970.1* | rs79851587 | A/C | 0.02 | 0.46 | 4.04E-08 | -0.11 | 0.13 | 18995 | No | 0.11 | 0.17 | 13379 | Yes | 0 | 0.88 | 285247 | Yes |
| *AC027119.1* | rs76221309 | T/C | 0.03 | 0.38 | 4.35E-08 | 0.07 | 0.24 | 18858 | Yes | 0.1 | 0.17 | 13099 | Yes | 0 | 0.97 | 285246 | Yes |
| *UBBP5* | rs117286162 | A/T | 0.02 | 0.44 | 4.68E-08 | 0.02 | 0.71 | 18995 | Yes | 0 | 0.96 | 13384 | Yes | -0.01 | 0.33 | 285248 | No |
| **Cavitated decay (Joint 2df test)** | | | | | | | | | | | | | | | | | |
| *RP11-527N22.1/KCNU1* | rs58016156 | G/A | 0.02 | - | 2.76E-08 | -0.52 | 0.34 | 1923 | - | - | - | - | - | - | - | - | - |
| **Clinical decay (GWAS Female stratum)** | | | | | | | | | | | | | | | | | |
| *RNA5SP388/NRXN3* | rs4899701 | T/A | 0.31 | -0.17 | 3.57E-08 | -0.05 | 0.22 | 19003 | No | 0.04 | 0.32 | 13385 | Yes | 0 | 0.71 | 285248 | Yes |
| **Cavitated decay status (binary) - (GWAS Female stratum)** | | | | | | | | | | | | | | | | | |
| *RP11-215I16.1* | rs200747282 | CA/C | 0.51 | 0.31 | 1.90E-08 | -0.01 | 0.82 | 4965 | - | 0.04 | 0.64 | 1865 | - | 0 | 0.73 | 284132 | - |
| **Cavitated decay status (binary) - (GWAS - Male stratum)** | | | | | | | | | | | | | | | | | |
| *RP11-933H2.4:NUDT16P1* | rs35487488 | T/C | 0.09 | -0.54 | 1.45E-08 | 0.1 | 0.24 | 18468 | No | -0.16 | 0.12 | 12685 | Yes | 0.01 | 0.09 | 285247 | No |

*Proxy SNP with R^2^>0.8 not found; **SNP is also genome-wide statistically significant for the joint test (main genetic effect + gene x environment effect);
Generalization: Nominal Significance + Directional consistency; GLIDE children study: Caries Yes/No trait.
Abbreviations: EA: effect allele; OA: Other allele; EAF - effect allele frequency; N - sample size; NA - Not applicable; SE: standard error; P: p-value; chr: Chromosome; pos: Position; 2df: 2-degree-of-freedom; DC: Directional consistency

**Appendix Table 12: Generalization of loci from ZOE-2.0 GWAS accounting for fluoride and sugar exposure (Approach 2 and 3) to GLIDE children and GLIDE adult studies.**

| **ZOE-2.0 study** | | | | | | **GLIDE children (Primary teeth)** | | | | **GLIDE children (Permanent teeth)** | | | | **GLIDE adults (DMFS and Dentures traits)** | | | |
| --- | --- | --- | --- | --- | --- | --- | --- | --- | --- | --- | --- | --- | --- | --- | --- | --- | --- |
| **Locus** | **rsid** | **EA/OA** | **EAF** | **beta** | **P** | **Beta** | **P** | **N** | **DC** | **Beta** | **P** | **N** | **DC** | **Beta** | **P** | **N** | **DC** |
| **Accounting for fluoride exposure** | | | | | | | | | | | | | | | | | |
| **Trait (Analysis): Cavitated decay (GWAS - Sub-optimal fluoride stratum)** | | | | | | | | | | | | | | | | | |
| *LEF1-AS1* | rs3913355 | G/A | 0.99 | -0.82 | 2.2E-08 | 0.17 | 0.49 | 4816 | Y | -0.69 | 0.32 | 426 | N | ** | ** | ** | ** |
| *SLC41A3*** | rs71327750 | T/G | 0.14 | 0.20 | 2.6E-08 | 0.08 | **0.02** | 18994 | Y | 0.01 | 0.76 | 13384 | Y | 0.00 | 0.83 | 285248 | Y |
| *NCOA2* | rs13256016 | A/G | 0.05 | 0.31 | 4.0E-08 | 0.01 | 0.91 | 18952 | Y | 0.08 | 0.10 | 13381 | Y | 0.01 | 0.18 | 285247 | Y |
| *AC110283.1* | rs138048415 | T/C | 0.02 | 0.65 | 4.8E-08 | 0.32 | 0.14 | 2454 | Y | -0.10 | 0.69 | 1628 | N | -0.01 | 0.23 | 273175 | N |
| **Cavitated decay (GWAS - Optimal fluoride stratum)** | | | | | | | | | | | | | | | | | |
| *RP1-62O9.3*** | rs650314 | C/A | 0.86 | -0.22 | 3.2E-09 | -0.04 | 0.38 | 18985 | N | -0.04 | 0.39 | 13380 | N | 0.00 | 0.51 | 285246 | N |
| *RP11-525K10.3*** | rs76985043 | C/G | 0.02 | 0.50 | 4.3E-09 | 0.00 | 0.94 | 18825 | N | -0.11 | 0.13 | 12997 | N | -0.01 | 0.23 | 285247 | N |
| *ZDHHC14*** | rs3861977 | T/A | 0.05 | 0.33 | 1.4E-08 | -0.01 | 0.81 | 13920 | Y | -0.05 | 0.34 | 13172 | Y | 0.00 | 0.35 | 285248 | N |
| **Clinical decay (GWAS - Optimal fluoride stratum)** | | | | | | | | | | | | | | | | | |
| *MACROD1/ RP11-21A7A.2* | rs12420136 | G/C | 0.04 | 0.39 | 2.5E-08 | -0.07 | 0.17 | 18873 | Y | -0.01 | 0.92 | 13350 | Y | 0.00 | 0.73 | 285248 | Y |
| **Accounting for Sugar exposure** | | | | | | | | | | | | | | | | | |
| **Cavitated decay (GWAS - High-sugar stratum)** | | | | | | | | | | | | | | | | | |
| *AC092635.1* | rs12052352 | T/G | 0.43 | 0.12 | 3.0E-08 | -0.01 | 0.64 | 18992 | N | -0.01 | 0.76 | 13385 | N | 0.00 | 0.32 | 285247 | Y |
| **Clinical decay (GWAS - High-sugar stratum)** | | | | | | | | | | | | | | | | | |
| *CTD-2555I5.1*** | rs11231965** | G/C | 0.11 | -0.21 | 7.8E-09 | -0.04 | 0.28 | 18982 | N | 0.02 | 0.52 | 13378 | Y | 0.00 | 0.98 | 285248 | Y |

*Could not find proxy with R2≥0.80; ⴕProxy SNPs not present in GLIDE datasets; **SNP is also genome-wide statistically significant for the joint test (main genetic effect + gene x environment effect); Generalization: Nominal Significance + Directional consistency; GLIDE children study: Caries Yes/No trait.
Abbreviations: EAF - effect allele frequency; N - sample size; NA - Not applicable; SE: standard error; chr: Chromosome; pos: Position; 2df: 2-degree-of-freedom; DC: Directional consistency

**Appendix Table 13: Look-ups in the ZOE 2.0 study results for the variants reported to be associated with caries in previous publications conducted in adults and children at a genome-wide significance level (5x10^-8^).**

| **Genome-wide statistically significant SNPs in published studies** | | | | | | **ZOE 2.0 results** | | | | |
| --- | --- | --- | --- | --- | --- | --- | --- | --- | --- | --- |
| **Locus** | **rsid** | **EA** | **Beta/ OR** | **P** | **N** | **EA** | **EAF** | **BETA** | **P** | **DC** |
| **Child Cohorts** | | | | | | | | | | |
| **Haworth et al. 2018** |  |  |  |  |  |  |  |  |  |  |
| *ALLC* | rs1594318 | C | -0.17 | 4.1E-08 | 16994 |  |  |  |  | - |
| *NEDD9* | rs7738851 | A | 0.25 | 1.6E-08 | 13353 | T | 0.40 | -0.01 | 0.66 | y |
| **Borgio et al. 2021** |  |  |  |  |  |  |  |  |  |  |
| *GRIN2B* | rs4764039 | C | 0.06 | 2.0E-08 | 111 | T | 0.12 | 0.05 | 0.11 | y |
| **Alotaibi et al. 2021** |  |  |  |  |  |  |  |  |  |  |
| *MIR3660* | rs80177293 | ND | -2.91 | 8.23E−10 | 1116 | C | 0.08 | -0.02 | 0.57 | - |
| *SUSD1/UGCG* | rs113021760 | ND | -2.67 | 3.36E−09 | 1116 | T | 0.04 | 0.07 | 0.11 | - |
| *ID4* | rs75833698 | ND | -2.17 | 4.93E−08 | 1116 |  |  |  |  | - |
| **Zeng et al. 2014** |  |  |  |  |  |  |  |  |  |  |
| *KPNA4* | rs17236529 | T | 2.81/1.78 | 2.0E-09 | 895 | T | 0.01 | -0.02 | 0.81 | n |
| **Adult cohorts** | | | | | | | | | | |
| **Shungin et al 2019** |  |  |  |  |  |  |  |  |  |  |
| *AMY1C* | rs72694438 | A | 0.02 | 2.3E-10 | 487822 | A | 0.15 | -0.01 | 0.72 | n |
| *KRTCAP2* | rs4971099 | A | -0.02 | 7.5E-15 | 487821 | G | 0.40 | 0.03 | 0.08 | y |
| *SYT14* | rs2046850 | T | -0.02 | 1.8E-08 | 487821 | T | 0.12 | 0.05 | 0.09 | n |
| *ITPKB* | rs3820640 | T | 0.02 | 2.7E-08 | 487822 | C | 0.07 | -0.02 | 0.66 | y |
| *KCNJ3* | rs2652452 | A | -0.02 | 3.3E-09 | 487823 | C | 0.37 | -0.01 | 0.73 | n |
| *ZNF804A* | rs263771 | A | 0.02 | 6.5E-09 | 487822 | A | 0.15 | 0.01 | 0.76 | y |
| *WNT10A* | rs121908120 | A | -0.08 | 2.0E-22 | 486339 |  |  |  |  |  |
| *ADCY3* | rs11676272 | A | -0.02 | 1.1E-08 | 487823 | G | 0.65 | -0.01 | 0.72 | n |
| *ALK* | rs80270335 | T | 0.03 | 2.1E-09 | 487821 | T | 0.04 | -0.05 | 0.28 | n |
| *FAM150B* | rs62106258 | T | 0.04 | 8.6E-12 | 487822 | C | 0.02 | -0.01 | 0.92 | y |
| *AAK1* | rs5831974 | D | -0.02 | 7.0E-10 | 486706 | TA | 0.68 | -0.01 | 0.49 | y |
| *STAG1* | rs61790808 | A | -0.02 | 9.1E-09 | 487820 | G | 0.45 | 0.03 | 0.16 | y |
| *KCNH8* | rs9831002 | T | -0.02 | 8.8E-10 | 487822 | G | 0.68 | 0.01 | 0.46 | y |
| *OPA1* | rs185566659 | A | 0.05 | 1.5E-09 | 487822 | A | 0.02 | 0.01 | 0.91 | y |
| *RARB* | rs7429279 | A | 0.02 | 1.3E-09 | 487822 | C | 0.51 | -0.01 | 0.53 | y |
| *RBM5* | 3:50135699DI | D | -0.02 | 4.8E-12 | 486705 |  |  |  |  |  |
| *EFNA5* | rs1352724 | A | -0.02 | 3.6E-08 | 487823 | A | 0.09 | -0.02 | 0.63 | y |
| *C5orf66* | rs1122171 | T | 0.04 | 2.8E-62 | 487823 | **T** | **0.55** | **0.04** | **0.01** | **y** |
| *CDH9* | rs55769264 | A | 0.02 | 2.8E-08 | 487822 | A | 0.21 | -0.02 | 0.47 | n |
| *FGF10* | rs1482698 | C | 0.02 | 1.5E-13 | 487823 | C | 0.24 | 0.02 | 0.35 | y |
| *HLA* | rs9366651 | T | -0.03 | 2.7E-28 | 487822 | T | 0.40 | 0.02 | 0.20 | n |
| *LOC157273* | rs898797 | T | 0.02 | 1.5E-08 | 487822 | T | 0.57 | 0.03 | 0.17 | y |
| *PBX3* | rs10987008 | A | 0.02 | 7.5E-14 | 487822 |  |  |  |  |  |
| *DMRTA1* | rs10811723 | A | -0.02 | 3.4E-11 | 487821 | G | 0.43 | 0.00 | 0.99 | y |
| *PRUNE2* | rs7852129 | A | -0.03 | 7.9E-11 | 483297 |  |  |  |  |  |
| *STFA1P* | rs7918807 | T | 0.02 | 3.6E-08 | 487821 | C | 0.61 | -0.02 | 0.20 | y |
| *P2RY2* | rs149467613 | A | -0.03 | 3.2E-08 | 487823 | A | 0.02 | 0.03 | 0.67 | n |
| *KLRAP1* | rs10772314 | A | -0.02 | 3.2E-08 | 487820 | A | 0.56 | 0.00 | 0.87 | n |
| *CA12* | rs72748935 | T | -0.03 | 1.3E-26 | 487820 | C | 0.23 | 0.02 | 0.33 | y |
| *NEO1* | rs6495046 | C | -0.02 | 3.5E-10 | 487821 | G | 0.78 | 0.00 | 0.98 | n |
| *CHRNA3* | rs10851907 | A | 0.02 | 1.0E-09 | 486706 | A | 0.38 | -0.01 | 0.44 | n |
| *RHCG* | rs2072693 | T | 0.01 | 4.9E-08 | 487821 | G | 0.70 | -0.01 | 0.47 | y |
| *TMEM219* | rs8054556 | A | 0.02 | 2.2E-09 | 487821 | A | 0.34 | 0.01 | 0.78 | y |
| *SALL1* | rs1108343 | T | 0.02 | 1.3E-08 | 487819 | C | 0.41 | -0.03 | 0.16 | y |
| *FOXL1* | rs10048146 | A | -0.03 | 5.2E-14 | 487820 | G | 0.15 | -0.01 | 0.74 | n |
| *NPEPPS* | rs3865314 | A | 0.02 | 1.5E-08 | 487822 | C | 0.39 | -0.02 | 0.18 | y |
| *HOXB-AS2* | rs9905793 | A | 0.03 | 6.5E-09 | 487820 | G | 0.80 | 0.00 | 0.97 | y |
| *KCNJ2* | rs34559440 | T | -0.02 | 1.1E-08 | 487822 | C | 0.26 | -0.01 | 0.49 | n |
| *LOC100499467* | rs7217268 | A | 0.02 | 1.5E-09 | 487821 | G | 0.75 | 0.00 | 0.86 | n |
| *BAHCC1* | rs57067187 | T | 0.02 | 6.9E-09 | 487821 | C | 0.17 | 0.03 | 0.20 | n |
| *MC4R* | rs28822480 | A | 0.02 | 7.1E-13 | 487821 | A | 0.22 | 0.02 | 0.28 | y |
| *CRLF1* | rs2238651 | T | 0.02 | 4.4E-08 | 487821 | T | 0.09 | -0.01 | 0.65 | n |
| *MAMSTR* | rs11672900 | A | -0.02 | 4.7E-14 | 487822 | G | 0.28 | 0.01 | 0.49 | y |
| *HAO1* | rs4816017 | A | -0.02 | 7.1E-09 | 487820 | A | 0.22 | -0.02 | 0.40 | y |
| *MTMR3* | rs140357883 | D | 0.02 | 2.3E-09 | 486706 |  |  |  |  |  |
| *FAM118A* | rs1569414 | T | -0.02 | 1.2E-11 | 487819 | G | 0.59 | 0.00 | 0.98 | n |
| *HDX* | rs5922945 | T | -0.02 | 1.6E-08 | 478256 |  |  |  |  | - |
| **Shaffer et al. 2013** |  |  |  |  |  |  |  |  |  |  |
| *AJAP1* | rs3896439 | NR | NR | 2.0E-08 | 920 | A | 0.11 | -0.01 | 0.86 | - |
| *LYZL2* | rs399593 | NR | NR | 9.0E-09 | 920 | T | 0.85 | 0.01 | 0.66 | - |

Abbreviations: EA: Effect allele, OR: Odds ratio, P: P-value, N: Sample size, EAF: Effect allele frequency, DC: Directional consistency of effect.

**Appendix Table 14: Annotation of novel loci identified from the Main Discovery analysis (Approach 1), the Joint test and the Stratum-specific GWAS (Approaches 2 and 3).**

| **(A) Main discovery analysis (Approach 1): Trait - Cavitated decay (d_3-6_mfs)** | | | | | | | | | | |
| --- | --- | --- | --- | --- | --- | --- | --- | --- | --- | --- |
| **Nearest Gene** | **Functional annotation** | **rsid** | **CADD^b^** | **RDB^c^** | **FATHMM-XF^d^** | **Promoter Histone markers^e^** | **Enhancer histone markers^e^** | **DNase^e^** | **Protein bound^e^** | **Motifs changes^e^** |
| *DLGAP1* | intronic | rs1442369 | 1.33 | 6 | 0.04, benign |  |  |  |  | Arid3a,PLZF,Sox |
| *SLC1A5* | intergenic | rs75906255 | 4.65 | 4 | 0.08, benign | SKIN, LNG, MUS | LNG, ESC, iPSC, ES-Deriv, BLD, STRM, FAT, MUS, SKIN, BRST, BRN, THYM, GI, PLCNT, KID, OVRY, LNG, CRVX, LIV, BON | LNG,ESDR,SKIN,BRN,MUS,GI,KID,LNG,OVRY,PLCNT,LIV |  | BDP1 |
| *RP11-527N22.1 / KCNU1* | intergenic | rs58016156 | 0.12 | 7 | 0.09, benign |  | BLD | THYM |  | HNF4,Znf143 |
| **(B) Accounting for Sex (Approach 2 and 3) - Trait /Sex Stratum** | | | | | | | | | | |
| **Cavitated decay (Male)** | | | | | | | | | | |
| *NDFIP1*** | intergenic | rs192232327 | 0.77 | 6 | 0.07, benign |  | LNG,ESDR,STRM,BRN,MUS,SKIN,BONE |  |  | GR_disc4, HNF4, Maf, NR4A, SREBP,Zbtb3 |
| *BNC2* | intergenic | rs191978580 | 5.50 | 7 | 0.07, benign |  |  |  |  | Ets,RREB-1,Rad21 |
| *RP11-856F16.2 / CWC15* | intergenic | rs74606067 | 0.51 | 7 | 0.10, benign |  | FAT, SKIN | IPSC |  |  |
| **Cavitated decay (Female)** | | | | | | | | | | |
| *NPAS3*** | intronic | rs2255032 | 11.70 | NA | 0.10, benign |  | MUS,SKIN,BRN,Gi,PANC | BRN |  | Evi-1,STAT |
| *AC016970.1 / MIR548A3* | intergenic | rs79851587 | 1.58 | 5 | 0.06, benign |  |  |  |  | Mef2 |
| *AC027119.1 / MIR4790* | intergenic | rs77865993 | 2.63 | 6 | 0.09, benign |  |  |  |  | 9 altered motifs |
| *LINC00438 / LOC642345* | intergenic | rs117286162 | 4.32 | 5 | 0.07, benign |  |  |  |  | Sox |
| **Clinical decay - Female** | | | | | | | | | | |
| *RN5S388 / NRXN3* | intergenic | rs4899701 | 0.071 | 7 | 0.13, benign |  |  |  |  |  |
| **Cavitated decay status (binary) - Female** | | | | | | | | | | |
| *RP11-480D4.1 / LOC729506* | intergenic | rs200747282 | 1.252 | NA | NA |  |  |  |  | AP-3,SP2 |
| **Cavitated decay status (binary) - Male** | | | | | | | | | | |
| *RP11-933H2.4 / NUDT16P1* | ncRNA_exonic | rs35487488 | 8.554 | 4 | 0.02, benign (high conf.) | 22 tissues | BLD, BRN, HRT | 28 tissues | YY1,POL2 | E2F,HNF4 |
| **(C) Accounting for Fluoride exposure (Approach 2 and 3) - Trait /Fluoride exposure stratum** | | | | | | | | | | |
| **Cavitated decay (Sub-optimal fluoride)** | | | | | | | | | | |
| *SLC41A3*** | intronic | rs71327750 | 0.63 | 5 | 0.02, benign (high conf.) |  |  |  |  | Nkx2 |
| *NCOA2* | intronic | rs13256016 | 3.33 | 7 | 0.03, benign |  | BRN, LIV | THYM |  | FAC1,TATA,YY1 |
| **Cavitated decay (Optimal fluoride)** | | | | | | | | | | |
| *RP1-62O9.3*** | ncRNA_intronic | rs650314 | 7.74 | 4 | 0.01, benign (high conf.) | CRVX | LNG,IPSC,ESDR,STRM,FAT,SKIN,BRST,BRN,MUS,HRT,GI,PLCNT,ADRL,SPLN,BLD,BONE | IPSC,SKIN,MUS,HRT,KID,LNG,PLCNT,CRVX,PANC,BRN | ERALPHA_A,TCF4,AP2ALPHA,AP2GAMMA, CEBPB | Mrg1::Hoxa9,NRSF,Pou2f2 |
| *RP11-525K10.3*** | ncRNA_intronic | rs76985043 | 1.16 | 7 | 0.05, benign |  |  |  |  |  |
| *ZDHHC14*** | intronic | rs3861977 | 2.64 | 4 | 0.03, benign (high conf.) | LNG,IPSC,ESDR,BLD,FAT,MUS,SKIN,BRST,BRN,GI,OVRY,ADRL,PLCNT,LIV,PANC,SPLN,CRVX,VAS,ESC,STRM,HRT,KID | ESC, BLD | ESDR,BLD,SKIN,THYM,BRN,GI,LNG |  | LBP-9,ZEB1 |
| **Clinical decay (Optimal fluoride)** | | | | | | | | | | |
| *MACROD1/ RP11-21A7A.2* | ncRNA_intronic | rs12420136 | 1.71 | 5 | 0.04, benign |  | *BRN, MUS* |  |  | HNF4,LF-A1,RAR,RXR::LXR,RXRA |
| **(D) Accounting for Sugar exposure (Approach 2 and 3) - Trait / Sugar exposure stratum** | | | | | | | | | | |
| **Cavitated decay / High sugar** | | | | | | | | | | |
| *AC092635.1* | upstream | rs12052352 | 6.45 | 5 | 0.06, benign | GI | LIV |  |  | *ATF3* |
| **Clinical decay / High sugar** | | | | | | | | | | |
| *CTD-2555I5.1*** | intergenic | rs11231965 | 0.36 | 7 | 0.16, benign |  | ESC, IPSC |  |  | Gm397 |

**SNP is also genome-wide statistically significant for the joint test (main genetic effect + gene x environment effect); Abbreviations: Chr - chromosome; EA: Effect allele; OA: Other allele; EAF - effect allele frequency; SNP - Single nucleotide polymorphisms; RDB: Regulome DB

b Scores the deleteriousness of the single nucleotide variants and INDELs in human genome (309).

c http://regulomedb.org; Likely to affect binding and linked to expression of a gene target (1a:eQTL + TF binding + matched TF motif + matched DNase Footprint + DNase peak, 1b:eQTL + TF binding + any motif + DNase Footprint + DNase peak, 1c:eQTL + TF binding + matched TF motif + DNase peak, 1d: eQTL + TF binding + any motif + DNase peak, 1e:eQTL + TF binding + matched TF motif; 1f:eQTL + TF binding / DNase peak); Likely to affect binding (2a: TF binding + matched TF motif + matched DNase Footprint + DNase peak, 2b:TF binding + any motif + DNase Footprint + DNase peak, 2c:TF binding + matched TF motif + DNase peak) (310).

d Based on Enhanced Accuracy in Predicting the Functional Consequences of Non-Coding and Coding Single Nucleotide Variants (FATHMM-XF) score (327).

e Promoter and Enhancer histone markers, DNAse, protein bound, and motif changes based on HaploReg v4.1 information (http://www.broadinstitute.org/mammals/haploreg/haploreg.php) that is derived from Roadmap Epigenomic Project and ENOCDE project (328).

f. Variant did not upload to FUMA, not found in 1000 Genomes data, with no information in HaploReg.

**Appendix Table 15: TAAR6 is the only genome-wide significant gene that emerged from gene-analysis using MAGMA (v1.6).**

| **Cavitated decay / GWAS Female-stratum** | | | | | | | | |
| --- | --- | --- | --- | --- | --- | --- | --- | --- |
| **GENE** | **CHR** | **START** | **STOP** | **NSNPS** | **NPARAM** | **N** | **P** | **SYMBOL** |
| ENSG00000146383 | 6 | 132891461 | 132892498 | 9 | 5 | 3002 | 1.1E-06 | *TAAR6* |

*Input SNPs were mapped to 18152 protein coding genes. Genome wide significance was defined at P = 0.05/18152 = 2.755e-6. Abbreviations: NSNPS: Number of SNPs in the gene NPARAM: Number of parameters used; N: Sample size, P: P-value

**Appendix Table 16: Significant gene sets with P_bon_<0.05 using the full distribution of SNP p-values.**

| **Gene Set** | **N genes** | **Beta** | **SE** | **P** | **P_bon_** |
| --- | --- | --- | --- | --- | --- |
| **Primary trait - Cavitated decay / p-difference (Sex-stratified)** | | | | | |
| GO_bp:go_regulation_of_adenylate_cyclase_activating_g_protein_coupled_receptor_signaling_pathway | 12 | 1.08 | 0.22 | 5.8E-07 | 9.0E-03 |
| **Secondary Traits** | | | | | |
| **Clinical decay - Main discovery** | | | | | |
| Curated_gene_sets:nikolsky_breast_cancer_1q32_amplicon | 13 | 1.38 | 0.30 | 2.5E-06 | 3.9E-02 |
| **Clinical decay / GWAS Female-stratum** | | | | | |
| GO_mf:go_taste_receptor_activity | 25 | 0.85 | 0.18 | 8.0E-07 | 1.2E-02 |
| **Cavitated decay status / p-difference (Sex-stratified)** | | | | | |
| Curated_gene_sets:pilon_klf1_targets_up | 455 | 0.19 | 0.04 | 4.9E-07 | 7.6E-03 |
| GO_bp:go_regulation_of_type_b_pancreatic_cell_development | 8 | 1.39 | 0.30 | 1.4E-06 | 2.1E-02 |
| GO_bp:go_type_b_pancreatic_cell_development | 20 | 0.86 | 0.18 | 1.4E-06 | 2.2E-02 |
| **Clinical decay / GWAS Sub-optimal fluoride-stratum** | | | | | |
| GO_bp:go_negative_regulation_of_potassium_ion_transport | 26 | 0.82 | 0.18 | 1.8E-06 | 2.8E-02 |
| **Clinical decay / p-difference (Sugar-stratified)** | | | | | |
| GO_bp:go_sequestering_of_metal_ion | 13 | 1.04 | 0.23 | 2.5E-06 | 3.8E-02 |
| **Clinical decay status / p-difference (Sugar-stratified)** | | | | | |
| GO_cc:go_interstitial_matrix | 12 | 1.19 | 0.26 | 2.7E-06 | 4.2E-02 |
| **Cavitated decay status / GWAS Low-Sugar-stratum** | | | | | |
| GO_bp:go_positive_regulation_of_rna_splicing | 32 | 0.61 | 0.13 | 2.5E-06 | 3.9E-02 |
| GO_cc:go_ino80_type_complex | 24 | 0.63 | 0.14 | 2.8E-06 | 4.4E-02 |

**Appendix Table 17: Enrichment of prioritized genes in curated genesets conducted in the GENE2FUNC process in FUMA.**

| **Primary Trait: Cavitated decay** | | | | | | | |
| --- | --- | --- | --- | --- | --- | --- | --- |
| **Trait / Analysis** | **Category** | **GeneSet** | **N genes** | **N overlap** | **p** | **adjP*** | **Genes** |
| Main Discovery | GWAScatalog | Male-pattern baldness | 250 | 16 | 6.9E-14 | 1.3E-10 | *FAR2:VIL1:USP37:RQCD1:PLCD4:ZNF142:BCS1L:RNF25:STK36:TTLL4:CYP27A1:PLCG1:ZHX3:LPIN3:EMILIN3:CHD6* |
|  | Positional_gene_sets | chr2q35 | 71 | 11 | 3.9E-14 | 1.2E-11 | *CTDSP1:VIL1:USP37:RQCD1:PLCD4:ZNF142:BCS1L:RNF25:STK36:TTLL4:CYP27A1* |
|  | Positional_gene_sets | chr20q12 | 7 | 5 | 5.1E-11 | 7.6E-09 | *PLCG1:ZHX3:LPIN3:EMILIN3:CHD6* |
| p-joint test for Sex-stratified | Positional_gene_sets | chr20q12 | 7 | 5 | 1.2E-10 | 3.6E-08 | *PLCG1:ZHX3:LPIN3:EMILIN3:CHD6* |
| p-difference test for Sex-stratified | GWAScatalog | Red vs. brown/black hair color | 18 | 5 | 8.1E-12 | 1.5E-08 | *FANCA:SPIRE2:TCF25:MC1R:TUBB3:TUBB3* |
| p-joint test for Sugar-stratified | Positional_gene_sets | chr20q12 | 7 | 5 | 1.6E-10 | 4.9E-08 | *PLCG1:ZHX3:LPIN3:EMILIN3:CHD6* |
| GWAS High-Sugar stratum | Positional_gene_sets | chr20q12 | 7 | 5 | 1.6E-11 | 4.7E-09 | *PLCG1:ZHX3:LPIN3:EMILIN3:CHD6* |
| GWAS Sub-optimal Fluoride stratum | GWAScatalog | Alzheimer's disease or fasting glucose levels (pleiotropy) | 45 | 12 | 3.6E-19 | 6.6E-16 | *MYBPC3:SPI1:SLC39A13:PSMC3:CELF1:FAM180B:C1QTNF4:MTCH2:AGBL2:FNBP4:NUP160:PTPRJ* |
|  | GWAScatalog | Loneliness (MTAG) | 67 | 11 | 4.1E-15 | 3.7E-12 | *MYBPC3:SPI1:SLC39A13:PSMC3:CELF1:FAM180B:C1QTNF4:MTCH2:AGBL2:FNBP4:NUP160* |
|  | GWAScatalog | Loneliness | 91 | 11 | 1.4E-13 | 8.4E-11 | *MYBPC3:SPI1:SLC39A13:PSMC3:CELF1:FAM180B:C1QTNF4:MTCH2:AGBL2:FNBP4:NUP160* |
|  | GWAScatalog | Global electrical heterogeneity phenotypes | 18 | 7 | 5.7E-13 | 2.6E-10 | *SPI1:SLC39A13:PSMC3:CELF1:AGBL2:FNBP4:NUP160* |
|  | GWAScatalog | Neuroticism | 138 | 11 | 1.4E-11 | 5.2E-09 | *MYBPC3:SPI1:SLC39A13:PSMC3:CELF1:FAM180B:C1QTNF4:MTCH2:AGBL2:FNBP4:NUP160* |
| **Secondary Traits** | | | | | | | |
| **Trait / Analysis** | **Category** | **GeneSet** | **N genes** | **N overlap** | **p** | **adjP** | **Genes** |
| **Clinical decay** | | | | | | | |
| Main discovery | Positional_gene_sets | chr20q12 | 7 | 5 | 9.6E-14 | 2.9E-11 | *PLCG1:ZHX3:LPIN3:EMILIN3:CHD6* |
| p-joint test for Sex-stratified | Positional_gene_sets | chr20q12 | 7 | 5 | 2.0E-13 | 5.9E-11 | *PLCG1:ZHX3:LPIN3:EMILIN3:CHD6* |
| p-difference test for Sex-stratified | Positional_gene_sets | chr2p14 | 18 | 5 | 1.3E-11 | 4.0E-09 | *PELI1:AFTPH:SERTAD2:CEP68:RAB1A* |
| GWAS Male stratum | Positional_gene_sets | chr8q23 | 16 | 5 | 3.1E-12 | 9.4E-10 | *NUDCD1:ENY2:PKHD1L1:EBAG9:SYBU* |
| p-joint test for Sugar-stratified | Positional_gene_sets | chr20q12 | 7 | 5 | 7.9E-14 | 2.4E-11 | *PLCG1:ZHX3:LPIN3:EMILIN3:CHD6* |
| GWAS High-Sugar stratum | Positional_gene_sets | chr20q12 | 7 | 6 | 1.4E-16 | 4.1E-14 | *TOP1:PLCG1:ZHX3:LPIN3:EMILIN3:CHD6* |
| GWAS Optimal-Fluoride stratum | GWAScatalog | Generalized epilepsy | 25 | 6 | 2.6E-13 | 4.7E-10 | *PNPO:COPZ2:NFE2L1:CBX1:SNX11:SKAP1* |
| **Cavitated decay status** | | | | | | | |
| p-difference test for Fluoride-stratified | GWAScatalog | Bipolar disorder | 590 | 12 | 7.5E-12 | 1.4E-08 | *MRPL11:PELI3:BBS1:ZDHHC24:CCS:RBM14:RBM4:RBM4B:SPTBN2:C11orf80:PC:C11orf86* |
|  | Chemical_and_Genetic_pertubation | NIKOLSKY_BREAST_CANCER_11Q12_Q14_AMPLICON | 156 | 14 | 5.8E-23 | 1.9E-19 | *MRPL11:PELI3:BBS1:ZDHHC24:CCS:RBM14:RBM4:RBM4B:SPTBN2:C11orf80:TMEM134:AIP:C11orf72:NDUFV1* |
|  | Positional_gene_sets | chr11q13 | 260 | 17 | 8.0E-26 | 2.4E-23 | *MRPL11:PELI3:BBS1:ZDHHC24:CCS:RBM14:RBM4:RBM14-RBM4:RBM4B:SPTBN2:C11orf80:PC:C11orf86:TMEM134:AIP:C11orf72:NDUFV1* |
|  | Curated_gene_sets | NIKOLSKY_BREAST_CANCER_11Q12_Q14_AMPLICON | 156 | 14 | 5.8E-23 | 3.2E-19 | *MRPL11:PELI3:BBS1:ZDHHC24:CCS:RBM14:RBM4:RBM4B:SPTBN2:C11orf80:TMEM134:AIP:C11orf72:NDUFV1* |
| **Clinical decay status** | | | | | | | |
| GWAS Female stratum | GWAScatalog | Mitochondrial DNA copy number | 26 | 8 | 2.6E-19 | 4.8E-16 | *CTNNA3:NR2F6:USHBP1:BABAM1:ANKLE1:ABHD8:MRPL34:DDA1* |

*This table only presents the results with the Adjusted p-value<5x10^-8^

**Appendix Table 18. Description and key findings of previous genome-wide association studies of dental caries carried out in children.**

| **Author /Year** | **N** | **Study Name** | **Age (years)** | **Race/Ethnicity*** | **Phenotype** | **Significant SNP (gene)** | **Suggestive SNP (gene)** |
| --- | --- | --- | --- | --- | --- | --- | --- |
| Shaffer et al., 2011 | 1,305 | COHRA, IFS, IHS | 3 to 12 | White | dft≥1 (Y/N) | None | rs2071862 (*TF1P11*), rs12191601 (*EPHA7*), rs6691598 (*ZMPSTE24*), rs110331093 (*MPPED2*) |
| Zeng et al, 2014 | 1,006 | COHRA, IFS | 3 to 12 | White | dfs count 1. Pit and fissure, 2. Smooth surface | rs17236529 (*KPNA4*), rs17236537 (*KPNA4*) | 92 loci |
| Haworth et al, 2018 | 19,003 | ALSPAC, YFS, GINI/ LISA, RAINE, GENEVA-Caries, COPSAC, PANIC, GenR, DNBC | 2.5 to 18 | European + 777 non-European ethnic groups | Affected vs caries-free | rs1594318 (*ALLC*) - primary | rs1594318 (*ALLC*), rs872877 (*ALLC*) |
| Alotaibi et al., 2021 | 1,116 | International study (US, Hungary, Argentina, Philippines, Guatemala) | 2 to 12 | Multi-ethnic | dft count | rs113021760 (*SUSD1*), rs16948495 (*DLX3, DLX4*), rs80177293 (*MIR3660*), rs75833698 (*ID4*) | rs75459295 (*SPTSSA*), chr9:35,753,170 (*CA9, TLN1*) |
| Borgio et al., 2021 | 111 | Saudi Arabia (Eastern province) | 2 to 6 | Arab | 76 cases (dmft >5); 35 controls (dmft = 0) | rs4764039C (*GRIN2B*); rs1065489G (*CFH*) | 6 loci |
| Orlova et al., 2022 | 2,974 | COHRA1, COHRA2, ALSPAC, IFS, IHS | Upto 5 | White (not defined in paper) | Affected vs caries-free | None | 7 loci |
| Ballantine et al, 2018 (Pilot) | 212 | ZOE | 2 to 4 | Multi-ethnic | dmfs≥1 (Y/N) | None | rs4690994 (Unknown), rs439888 (*CLDN14*) |
| Orlova et al., 2019 (Pilot) | 96 | COHRA1 | 3 to 12 | African American | dft/dfs count | None | 42 loci |

*As defined in the publication. Abbreviations: N: Sample size; COHRA:Center for Oral Health Research in Appalachia; IFS: Iowa Fluoride Study; IHS: Iowa Head Start Study; DRDR1: Dental Registry and DNA Repository cohort phase 1; DRDR2: Dental Registry and DNA Repository cohort phase 2; ARIC: Atherosclerosis Risk in Community cohort; HPFS: Health Professionals Follow-Up Study cohort; GEIRS: Genetic, Environment and Health Initiative Research study; ZOE: Zero-Out Early Childhood Caries; DFT: Decayed and filled teeth (permanent dentition); DMFS: Decayed, Missing, Filled surfaces; DFS: Decayed and Filled surfaces; dft: decayed and filled teeth (primary dentition); dfs: decayed and filled surfaces (primary dentition);
